# Redefining Synovial Pathotypes in Psoriatic Arthritis through Multi-Modal Histology and Machine Learning (The PSYNOVIUM Study)

**DOI:** 10.64898/2026.11.06.26367442

**Authors:** Ryan Malcolm Hum, Maria Christofi, Samantha L Smith, Yvonne Tan, Rudresh Shukla, Lysette Michele Marshall, Megan Smith, Alec Williams, Gisela Orozco, Sebastien Viatte, Maya Buch, Anne Barton, Pauline Ho, Paul Martin

## Abstract

**Background:** Synovial inflammation in psoriatic arthritis (PsA) lacks a validated histological classification framework. Current approaches apply rheumatoid arthritis (RA) synovial pathotypes to PsA, which overlooks immunobiological differences between the diseases and limits the predictive power of tissue stratification. We aimed to define data-driven, PsA-specific synovial pathotypes to facilitate tissue-driven precision medicine.

**Methods:** In this cross-sectional study, ultrasound-guided synovial biopsies were obtained from 61 patients with PsA (n=22), seropositive RA (n=23), seronegative RA (n=10), and other inflammatory arthritides (n=6). Sections underwent multi-modal staining, including H&E, picrosirius red, and immunohisto-chemistry for CD3, CD20, CD68, CD138, and CD31. Whole-slide digital image analysis and automated pixel classifiers quantified cellular and stromal metrics. Unsupervised hierarchical clustering identified intrinsic PsA pathotypes, which were correlated with clinical disease activity, radiographic osteoarthritis (OA), and longitudinal treatment responses.

**Findings:** While a subset of PsA patients shared an inflammatory signature with seropositive RA, unsupervised clustering of the PsA cohort revealed three distinct, novel pathotypes: ‘Dense Myeloid’ (macrophage-driven), ‘Fibro-Stromal’ (fibrosis and pauci-immune), and ‘Angio-Lymphoid’ (hypervascular with significant CD138^+^ plasma cell infiltration). Pathotypes were independent of treatment and concurrent radiographic knee OA. The Fibro-Stromal pathotype correlated with significantly higher cutaneous psoriasis severity. Longitudinally, patients with the Fibro-Stromal pathotype exhibited a trend toward lower treatment response (44%) following standard interventions (methotrexate or TNF inhibitors) compared to the Dense Myeloid (50%) and Angio-Lymphoid (80%) pathotypes.

**Interpretation:** We present the first computationally derived synovial pathotypes specific to PsA, revealing distinct plasma cell and fibrotic architectures not captured by RA-centric models. To enable clinical translation, we distilled these findings into a pragmatic, four-marker diagnostic decision tree (CD31, CD68, CD138, and picrosirius red) designed for routine histopathology workflows, providing a staging ground for biomarker-stratified clinical trials in PsA.

**Funding:** Funding from the NIHR Biomedical Research Centre: Manchester, Arthritis UK, the Wellcome Trust, Brit-PACT, the National Psoriasis Foundation, and the Medical Research Council.

**Research in context:** *Evidence before this study:* The potential clinical utility of tissue-based patient stratification is well established in rheumatoid arthritis (RA), where baseline synovial pathotypes (lympho-myeloid, diffuse-myeloid, and pauciimmune) have guided targeted biologic therapy in landmark precision medicine trials. However, psoriatic arthritis (PsA) exhibits a fundamentally distinct synovial microenvironment, characterised by unique vascular architecture, altered stromal fibroblast phenotypes, and distinct immune infiltrates intertwined with the IL-23/IL-17 cytokine axis. Consequently, attempting to classify PsA synovial tissue using established RA-derived scoring systems risks misrepresenting intrinsic disease biology. There is currently no validated, disease-specific histological classification system for PsA, which forces therapeutic decision-making to rely on an empirical, trial-and-error approach rather than underlying target tissue pathobiology.

*Added value of this study:* To our knowledge, this is the first study to define data-driven, computationally derived synovial pathotypes entirely specific to PsA. By leveraging multi-modal digital pathology and unsupervised machine learning, we deconstructed the PsA synovium into three biologically distinct pathotypes: Dense Myeloid, Fibro-Stromal, and Angio-Lymphoid. We demonstrated that the highly inflammatory PsA microenvironment is distinctly driven by CD138^+^ plasma cells rather than the CD20^+^ B cells typically observed in RA. Furthermore, we identified a striking tissue-level divergence: patients with the highly fibrotic, pauci-immune Fibro-Stromal pathotype exhibited the most severe cutaneous psoriasis and demonstrated a trend toward higher failure rates on standard therapies, independent of concurrent osteoarthritis. Crucially, we translated these high-dimensional computational findings into a pragmatic, four-marker diagnostic decision tree (incorporating CD31, CD68, CD138, and picrosirius red).

*Implications of all the available evidence:* This study establishes that PsA synovial pathology requires a dedicated, disease-specific classification framework. The proposed diagnostic decision tree can be integrated into routine clinical histopathology workflows without the immediate need for advanced spatial or single-cell transcriptomics. Furthermore, the correlation between the Fibro-Stromal pathotype, severe skin disease, and poor joint response to standard therapies provides a biological rationale for stratifying this specific subgroup towards early IL-17 or IL-23 inhibitor intervention alongside conventional synthetic disease-modifying antirheumatic drugs. Ultimately, these novel PsA pathotypes provide the vital staging ground required to design and execute biomarker-stratified, tissue-driven clinical trials, paving the way for precision medicine in PsA.

## Introduction

Psoriatic arthritis (PsA) is a prevalent inflammatory arthritis characterised by heterogeneous joint involvement and cutaneous psoriasis.^1, 2^ While the therapeutic armamentarium for PsA has expanded significantly to include biologic and targeted synthetic disease-modifying antirheumatic drugs (b/tsDMARDs), treatment selection remains largely empirical. Approximately half of patients fail to achieve minimal disease activity,^3^ and therapeutic decisions in clinical practice are primarily driven by trial-and-error, drug costs, or clinical contraindications rather than underlying disease mechanisms.^4, 5^ Although hallmark features of PsA synovitis, such as pronounced, tortuous neoangiogenesis,^6, 7^ lining layer hyperplasia,^8, 9^ and specific immune cell infiltrates intertwined with the IL-23/IL-17 cytokine axis,^2, 10–19^ are well documented, PsA critically lacks a standardised, validated histological classification framework. This absence prevents the stratification of patients based on synovial pathobiology, representing a major bottleneck to precision medicine.

The potential clinical utility of tissue-based patient stratification is already well established in rheumatoid arthritis (RA). In RA, the synovial microenvironment can be reliably classified into three distinct pathotypes: lympho-myeloid, diffuse-myeloid, and pauci-immune.^20^ These signatures correlate with clinical disease activity, radiographic progression, and treatment response.^20^ More recently, similar pathotypic structures have been identified in osteoarthritis (OA) and systemic sclerosis synovium.^21–23^ Crucially, landmark biopsy-driven clinical trials, such as R4RA and STRAP,^24–26^ have validated this paradigm by demonstrating that baseline RA pathotypes can guide targeted biologic therapy. This precedent establishes that integrating synovial histopathology into routine clinical decision-making is both viable and potentially impactful.

However, applying RA-derived classification systems directly to PsA overlooks the fundamental immunopathological and genetic divergence between the two diseases.^2, 27, 28^ The PsA synovial microenvironment exhibits a highly distinct cellular milieu, defined by the enrichment of IL-17-producing T cells,^29^ specific myeloid populations,^13, 14, 17, 30–33^ and mast cells^8, 18^ operating alongside unique fibroblast phenotypes^31, 34–36^ thought to mediate aberrant osteoproliferation and erosive damage. Furthermore, the defining architectural features of PsA (notably its pronounced neoangiogenesis and distinct spatial distribution of sublining infiltrates) are inadequately captured by RA-centric scoring algorithms.^2^ Consequently, attempting to assign PsA synovial tissue into RA-derived pathotypes risks misrepresenting intrinsic disease biology and diluting the predictive power of tissue stratification. This highlights an urgent, unmet need to develop data-driven synovial pathotypes specific to PsA.

To address this gap, this study aimed to define clinically meaningful, data-driven PsA synovial pathotypes using multi-modal histology and machine learning, and to subsequently construct a clinically translatable diagnostic decision tree. Moving beyond traditional, subjective, and semi-quantitative manual scoring, we leveraged whole-slide digital pathology to extract objective cellular and stromal metrics from the synovial microenvironment. By applying unsupervised machine learning techniques to these datasets, we sought to uncover intrinsic, biologically meaningful histological subgroups within PsA and contrast their features with seropositive and seronegative RA. Finally, to ensure clinical utility, we distilled these computationally derived signatures into a simplified, accessible immunohisto-chemistry (IHC) scoring tool. Designed for seamless implementation into routine pathology workflows, this novel, disease-specific classification system provides the critical framework required to facilitate future biomarker-stratified, tissue-driven clinical trials in PsA.

## Methods

### Patient Recruitment and Synovial Biopsy

This cross-sectional, observational study evaluated inflamed synovial tissue from patients with PsA (fulfilling CASPAR criteria) and seropositive (defined strictly by anti-citrullinated protein antibody [ACPA] positivity, independent of rheumatoid factor status) or seronegative (ACPA negative, independent of rheumatoid factor status) RA (fulfilling 2010 ACR/EULAR criteria). Following informed written consent, patients with ultrasound-detected joint inflammation (grey-scale *≥*2 and power Doppler *≥*1) underwent ultrasound-guided, minimally invasive synovial needle core biopsy. The anatomical site of the biopsied joint was recorded for all patients. Baseline clinical metrics including psoriasis body surface area (BSA), joint counts, global assessments, and acute phase reactants were captured at the time of biopsy. To account for potential confounding from secondary degenerative changes, particularly given the high proportion of knee biopsies, concurrent radiographic osteoarthritis (OA) severity at the biopsied joint was evaluated from pre-biopsy imaging and graded as none, mild, moderate, or severe.^37^ Additionally, longitudinal clinical trajectories were tracked from the date of biopsy to the most recent clinical follow-up to evaluate post-biopsy treatment changes and therapeutic responses. Therapeutic efficacy was classified as a good/moderate versus poor response utilising EULAR response criteria (derived from DAS28 scores) for RA patients,^38^ and the Psoriatic Arthritis Response Criteria (PsARC) for PsA patients.^39^

### Histological and Immunohistochemical Staining

Synovial biopsy cores were formalin-fixed and paraffin-embedded (FFPE). To comprehensively capture synovial inflammation, stromal fibrosis, and vascularity, serial 5µm sections underwent a multi-modal staining strategy. General morphological assessment was performed using Haematoxylin and Eosin (H&E) staining. Picrosirius red with a fast green counterstain was utilised to specifically identify and quantify the collagenous extracellular matrix. The immune and vascular microenvironments were characterised via automated immunohistochemistry (IHC) using a validated panel of lineage markers: CD3 (T cells), CD20 (B cells), CD68 (macrophages), CD138 (plasma cells), and CD31 (endothelial cells).

### Digital Image Analysis and Pathotyping

High-resolution whole-slide images were analysed in a blinded manner using QuPath software (v0.6.0). Standardised Krenn synovitis scores and digital cellularity (cells/mm^2^) were derived from H&E sections. Established RA-derived synovial pathotypes (lympho-myeloid, diffuse-myeloid, and pauciimmune) were assigned based on conventional semi-quantitative (0–3) scoring of the IHC lineage markers (Supplementary Figures 1 and 2).^20^ To validate these scores and generate robust continuous variables for cross-disease comparison, automated positive cell detection and high-resolution Random Trees pixel classifiers were trained and deployed across the entire cohort. This enabled the objective digital quantification of absolute positive cell percentages, CD31^+^ vascular area fraction, and proportional fibrotic burden.

### Unsupervised Clustering for Novel Pathotype Discovery

To identify novel, data-driven synovial pathotypes in PsA, unsupervised agglomerative hierarchical clustering (Ward’s linkage, Euclidean distance) was employed. An input matrix was constructed incorporating cellularity, fibrosis, vascularity, and the four quantitative IHC metrics. The complete cohort and the lymphomyeloid subset were clustered using k-means clustering to explore histological overlap between diagnoses, with elbow plots and silhouette scores used to evaluate the number of clusters. For PsA, three clusters were selected because three pathotypes were more informative than two or five and are analogous to established RA pathotypes.^20^ To facilitate the translation of these computationally derived pathotypes into routine clinical histopathological workflows, a diagnostic decision tree algorithm was subsequently developed. Marker selection for this algorithm was driven by cluster discrimination capacity, and practical alignment with existing four-marker plus H&E RA pathotyping protocols.

### Statistical Analysis

Statistical analyses were performed using Python (v3.12). Continuous variables were compared across clinical diagnoses and pathotypes using the Kruskal-Wallis or Mann-Whitney U test, as appropriate. P values were adjusted for multiple comparisons using the Benjamini-Hochberg false discovery rate (FDR) method where stated. Correlations between continuous digital pathology metrics and clinical disease activity scores were assessed using Spearman’s rank correlation. Receiver operating characteristic (ROC) curve analysis was utilised to evaluate the diagnostic utility of specific cellular markers in differentiating PsA from RA. Categorical variables, including radiographic OA severity and longitudinal treatment responses, were visualised using stacked bar charts and cross-tabulations. A p-value of <0.05 was considered statistically significant.

### Supplementary methods

Detailed patient recruitment protocols, ethical approvals, the standardised biopsy procedure, comprehensive tissue processing steps, staining hierarchies, specific antibody clones, and hyperparameter settings for machine learning algorithms are provided in the Supplementary Methods.

## Results

### Synovial tissue cohort and baseline histological characterisation

Synovial tissue samples were obtained from 61 patients, predominantly comprising individuals with PsA (n=22) and seropositive RA (n=23) (Table 1, Figure 1a). Inflammatory immunohistochemical profiles enabled the assignment of established RA-derived synovial pathotypes for 60 patients (where 1 patient had insufficient tissue to enable full profiling).^20^ Based on Krenn synovitis score and grade profiling (Figure 1e, Supplementary Figure 3), 49% of the cohort exhibited high-grade synovitis (Krenn score 5–9, n=30/61), 43% had low-grade synovitis (Krenn score 2–4, n=26/61), and 8% had no synovitis (Krenn score 0–1, n=5/61), with comparable distributions of scores and grades across diagnoses (Figures 1b and 1c, Supplementary Table 1).

**Figure 1.**
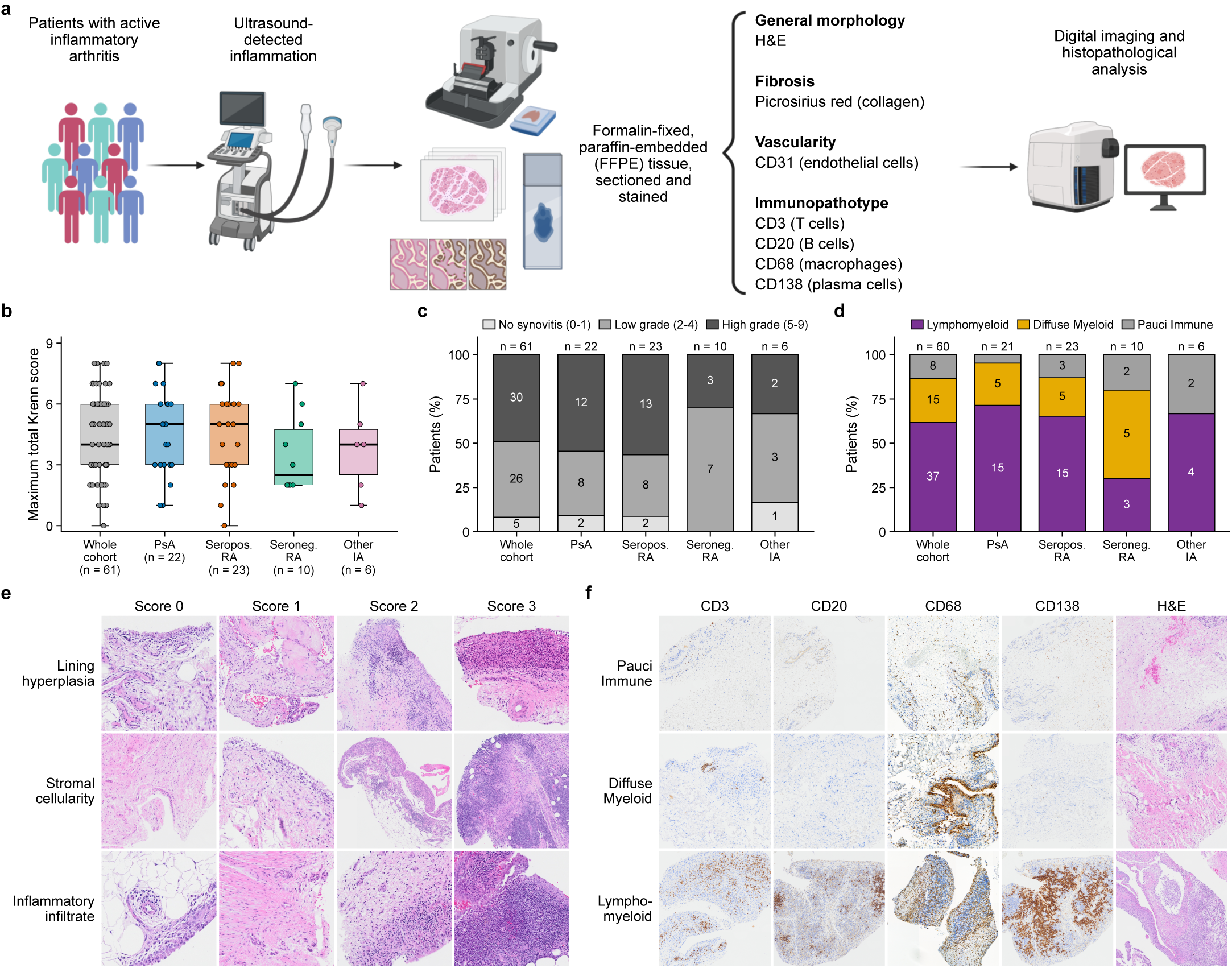
Overview of the multi-modal histology pipeline and cohort Krenn synovitis and traditional pathotype classification by disease which reveals overlap and similarities between PsA and RA. (a) Overview of patient recruitment and synovial tissue sample processing pipeline and histological profiling, created with BioRender. (b) Krenn synovitis score by diagnosis. (c) Krenn synovitis grade by diagnosis. (d) RA synovial pathotypes by diagnosis. (e) Representative synovial tissue images of Krenn domain scores from 0 to 3. (f) Representative synovial tissue images from each RA synovial pathotype in PsA. Images in (e) and (f) are shown at different magnifications.

**Table 1:** Cohort demographic and clinical characteristics.

|  | **Entire cohort (n=61)** | **PsA (n=22)** | **Seropositive RA (n=23)** | **Seronegative RA (n=10)** | **Other IA (n=6)** | **Missing (n [%])** | **p-value<sup>\*</sup>** |
|---|---|---|---|---|---|---|---|
| **Demographics** |  |  |  |  |  |  |  |
| Age, years (median [IQR]) | 52.0 (35.0–64.0) | 46.0 (32.0–66.8) | 43.0 (38.5–61.0) | 58.5 (56.2–63.2) | 46.0 (36.2–61.8) | 0 (0.0%) | 0.593 |
| Female sex (n [%]) | 37 (60.7%) | 9 (40.9%) | 16 (69.6%) | 7 (70.0%) | 5 (83.3%) | 0 (0.0%) | 0.102 |
| Disease duration, years (median [IQR]) | 3.5 (0.0–14.2) | 6.0 (0.2–15.0) | 2.5 (0.0–11.8) | 3.5 (1.0–20.0) | 11.5 (0.5–25.5) | 1 (1.6%) | 0.383 |
| Body mass index, kg/m<sup>2</sup> (median [IQR]) | 28.1 (24.0–31.6) | 30.2 (24.7–34.7) | 27.8 (23.4–31.3) | 27.6 (24.3–32.1) | 24.1 (23.8–26.0) | 0 (0.0%) | 0.137 |
| Ethnicity (n [%]) |  |  |  |  |  | 1 (1.6%) | – |
| White | 48 (78.7%) | 20 (90.9%) | 15 (65.2%) | 9 (90.0%) | 4 (66.7%) | – | – |
| Mixed | 1 (1.6%) | 0 (0.0%) | 0 (0.0%) | 0 (0.0%) | 1 (16.7%) | – | – |
| Black | 4 (6.6%) | 0 (0.0%) | 3 (13.0%) | 1 (10.0%) | 0 (0.0%) | – | – |
| Asian | 7 (11.5%) | 2 (9.1%) | 4 (17.4%) | 0 (0.0%) | 1 (16.7%) | – | – |
| Unknown | 1 (1.6%) | 0 (0.0%) | 1 (4.3%) | 0 (0.0%) | 0 (0.0%) | – | – |
| Smoking (n [%]) |  |  |  |  |  | 3 (4.9%) | 0.160 |
| Never | 35 (60.3%) | 16 (76.2%) | 10 (47.6%) | 3 (30.0%) | 6 (100.0%) | – | – |
| Current | 12 (20.7%) | 2 (9.5%) | 5 (23.8%) | 5 (50.0%) | 0 (0.0%) | – | – |
| Ex | 11 (19.0%) | 3 (14.3%) | 6 (28.6%) | 2 (20.0%) | 0 (0.0%) | – | – |
| **Serology and HLA-B27** |  |  |  |  |  |  |  |
| HLA-B27 positive (n [%])<sup>†</sup> | – | 4 (26.7%) | – | – | – | 43 (70.5%) | – |
| Anti-CCP positive (n [%]) | 23 (39.0%) | 0 (0.0%) | 23 (100.0%) | 0 (0.0%) | 0 (0.0%) | 2 (3.3%) | – |
| **Treatment** |  |  |  |  |  |  |  |
| Treatment naive (n [%]) | 19 (31.1%) | 5 (22.7%) | 10 (43.5%) | 3 (30.0%) | 1 (16.7%) | 0 (0.0%) | 0.246 |
| Current csDMARDs, number (median [IQR]) | 1.0 (0.0–1.0) | 0.5 (0.0–1.0) | 0.0 (0.0–1.0) | 0.5 (0.0–1.0) | 1.0 (1.0–1.0) | 0 (0.0%) | 0.889 |
| On methotrexate (n [%]) | 20 (32.8%) | 6 (27.3%) | 7 (30.4%) | 5 (50.0%) | 2 (33.3%) | 0 (0.0%) | 1.000 |
| On sulfasalazine (n [%]) | 6 (9.8%) | 4 (18.2%) | 1 (4.3%) | 1 (10.0%) | 0 (0.0%) | 0 (0.0%) | 0.317 |
| On hydroxychloroquine (n [%]) | 11 (18.0%) | 2 (9.1%) | 6 (26.1%) | 1 (10.0%) | 2 (33.3%) | 0 (0.0%) | 0.271 |
| Biologics ever, number (median [IQR]) | 0.0 (0.0–3.0) | 0.0 (0.0–2.8) | 0.0 (0.0–3.0) | 0.0 (0.0–2.5) | 0.0 (0.0–1.5) | 0 (0.0%) | 0.901 |
| Biologic experienced (n [%]) | 25 (41.0%) | 9 (40.9%) | 10 (43.5%) | 4 (40.0%) | 2 (33.3%) | 0 (0.0%) | 1.000 |
| Current biologic or targeted synthetic DMARD (n [%])<sup>‡</sup> | 23 (37.7%) | 8 (36.4%) | 9 (39.1%) | 4 (40.0%) | 2 (33.3%) | 0 (0.0%) | – |
| Filgotinib | 2 (8.7%) | 0 (0.0%) | 1 (11.1%) | 1 (25.0%) | 0 (0.0%) | – | – |
| Rituximab | 4 (17.4%) | 0 (0.0%) | 2 (22.2%) | 1 (25.0%) | 1 (50.0%) | – | – |
| Secukinumab | 2 (8.7%) | 2 (25.0%) | 0 (0.0%) | 0 (0.0%) | 0 (0.0%) | – | – |
| Upadacitinib | 1 (4.3%) | 0 (0.0%) | 1 (11.1%) | 0 (0.0%) | 0 (0.0%) | – | – |
| Baricitinib | 3 (13.0%) | 0 (0.0%) | 3 (33.3%) | 0 (0.0%) | 0 (0.0%) | – | – |
| Adalimumab | 3 (13.0%) | 2 (25.0%) | 1 (11.1%) | 0 (0.0%) | 0 (0.0%) | – | – |
| Tocilizumab | 3 (13.0%) | 1 (12.5%) | 1 (11.1%) | 1 (25.0%) | 0 (0.0%) | – | – |
| Sarilumab | 1 (4.3%) | 1 (12.5%) | 0 (0.0%) | 0 (0.0%) | 0 (0.0%) | – | – |
| Abatacept | 1 (4.3%) | 0 (0.0%) | 0 (0.0%) | 1 (25.0%) | 0 (0.0%) | – | – |
| Adalimumab, Apremilast | 1 (4.3%) | 1 (12.5%) | 0 (0.0%) | 0 (0.0%) | 0 (0.0%) | – | – |
| Etanercept | 1 (4.3%) | 0 (0.0%) | 0 (0.0%) | 0 (0.0%) | 1 (50.0%) | – | – |
| Guselkumab | 1 (4.3%) | 1 (12.5%) | 0 (0.0%) | 0 (0.0%) | 0 (0.0%) | – | – |
| TNF inhibitor exposure (n [%]) |  |  |  |  |  | 0 (0.0%) | 0.454 |
| TNF inhibitor naive | 37 (60.7%) | 13 (59.1%) | 13 (56.5%) | 7 (70.0%) | 4 (66.7%) | – | – |
| Previous TNF inhibitor | 19 (31.1%) | 6 (27.3%) | 9 (39.1%) | 3 (30.0%) | 1 (16.7%) | – | – |
| On TNF inhibitor | 5 (8.2%) | 3 (13.6%) | 1 (4.3%) | 0 (0.0%) | 1 (16.7%) | – | – |
| **Disease activity<sup>§</sup>** |  |  |  |  |  |  |  |
| ESR, mm/h (median [IQR]) | 29.5 (10.8–47.5) | 24.5 (15.2–44.8) | 38.0 (12.5–65.2) | 30.5 (22.2–35.8) | 12.0 (6.8–23.2) | 1 (1.6%) | 0.318 |
| CRP, mg/L (median [IQR]) | 12.5 (5.8–28.5) | 11.0 (6.0–15.8) | 23.0 (10.8–41.5) | 10.5 (4.5–19.2) | 7.0 (1.5–15.5) | 1 (1.6%) | 0.050 |
| **Psoriatic Arthritis Specific Measures of Disease Activity** |  |  |  |  |  |  |  |
| Tender joint count (68) (median [IQR]) | – | 6.5 (3.2–12.8) | – | – | 2.5 (1.8–3.2) [n=2/6] | – | – |
| Swollen joint count (66) (median [IQR]) | – | 6.5 (3.0–13.2) | – | – | 2.5 (1.8–3.2) [n=2/6] | – | – |
| Patient global (1–5) (median [IQR]) | – | 4.0 (3.0–5.0) | – | – | 3.0 (3.0–3.0) [n=1/6] | – | – |
| Physician global (1–5) (median [IQR]) | – | 4.0 (3.0–4.0) | – | – | 3.0 (3.0–3.0) [n=1/6] | – | – |
| Enthesitis (6 sites) (median [IQR]) | – | 0.0 (0.0–2.0) [n=20/22] | – | – | – | – | – |
| Dactylitis (10 digits) (median [IQR]) | – | 0.0 (0.0–0.2) [n=20/22] | – | – | – | – | – |
| Nail involvement (n [%]) | – | 9 (42.9%) [n=21/22] | – | – | – | – | – |
| PASI (median [IQR]) | – | 0.0 (0.0–2.7) [n=17/22] | – | – | – | – | – |
| Body surface area, % (median [IQR]) | – | 1.0 (0.0–2.8) | – | – | – | – | – |
| **Rheumatoid Arthritis Specific Measures of Disease Activity** |  |  |  |  |  |  |  |
| Tender joint count (28) (median [IQR]) | – | – | 8.0 (6.0–14.5) | 11.5 (9.0–21.5) | 2.5 (1.0–7.2) [n=4/6] | – | – |
| Swollen joint count (28) (median [IQR]) | – | – | 7.0 (4.5–14.0) | 6.0 (3.5–16.8) | 2.5 (1.0–4.2) [n=4/6] | – | – |
| Patient global (0–100) (median [IQR]) | – | – | 80.0 (70.0–97.5) | 85.0 (80.0–90.0) [n=9/10] | 100.0 (100.0–100.0) [n=1/6] | – | – |

<sup>\*</sup>*P values compare PsA with seropositive RA, by Mann-Whitney U test for continuous variables and chi-squared test for categorical variables.*
<sup>†</sup>*Percentages are of patients tested. HLA-B27 was tested selectively: 15/22 PsA, 0/23 Seropositive RA, 1/10 Seronegative RA, 2/6 Other IA.*
<sup>‡</sup>*Percentages in the subrows are of patients on a biologic or targeted synthetic DMARD in that group.*
<sup>§</sup>*Disease-activity instruments differed by diagnosis. “–” means not recorded in that group. [n=] is the number of patients with a value, where fewer than the whole group.*
*csDMARD, conventional synthetic disease-modifying antirheumatic drug; DMARD, disease-modifying antirheumatic drug; CRP, C-reactive protein; ESR, erythrocyte sedimentation rate; IA, inflammatory arthritis; IQR, interquartile range; PASI, Psoriasis Area and Severity Index; PsA, psoriatic arthritis; RA, rheumatoid arthritis; TNF, tumour necrosis factor.*

### High-grade synovitis aligns with a lymphomyeloid pathotype in PsA and seropositive RA

The lymphomyeloid pathotype was the most frequently assigned signature across the entire cohort (62%, n=37/60; Figures 1d and 1f, Supplementary Figure 4). The proportion of patients assigned a lymphomyeloid pathotype was comparable between the PsA and seropositive RA cohorts (71%, n=15/21 vs 65%, n=15/23, respectively). Integrating pathotype assignments with Krenn synovitis grading revealed that most patients with high-grade synovitis (Krenn score 5–9) exhibited a lymphomyeloid pathotype (93%, n=28/30). Notably, 92% of PsA patients (n=11/12) and 100% of seropositive RA patients (n=13/13) with high-grade synovitis were assigned this pathotype (Supplementary Figure 5b).

### Macrophage (CD68) expression differentiates PsA from seronegative RA

Digital pathology assessments and consensus semi-quantitative scoring demonstrated high concordance across all histological markers (Supplementary Figures 6a–h and 7a–d). Comparison of digital pathology scores for individual histological markers by diagnosis revealed similar expression levels of CD3, CD20, CD138, and overall cellularity across the different arthritides (Figures 2a, 2b, 2d and 3a, Supplementary Figure 7e, Supplementary Table 2). However, CD68 expression was significantly higher in patients with PsA compared to those with seronegative RA (19.0% vs 9.4% positivity, FDR p=0.007, respectively) (Figure 2c, Supplementary Figure 7e). Receiver operating characteristic (ROC) curve analysis utilising CD68 as a classifier to differentiate PsA from seronegative RA yielded an area under the curve (AUC) of 0.85 (95% CI 0.68–0.98, p=0.002). A threshold of 11.6% positive cell detection provided a sensitivity of 81% and a specificity of 80% (Figure 2e). CD68 did not differentiate PsA from seropositive RA (AUC 0.57, 95% CI 0.38–0.74, p=0.466) (Figure 2f).

**Figure 2.**
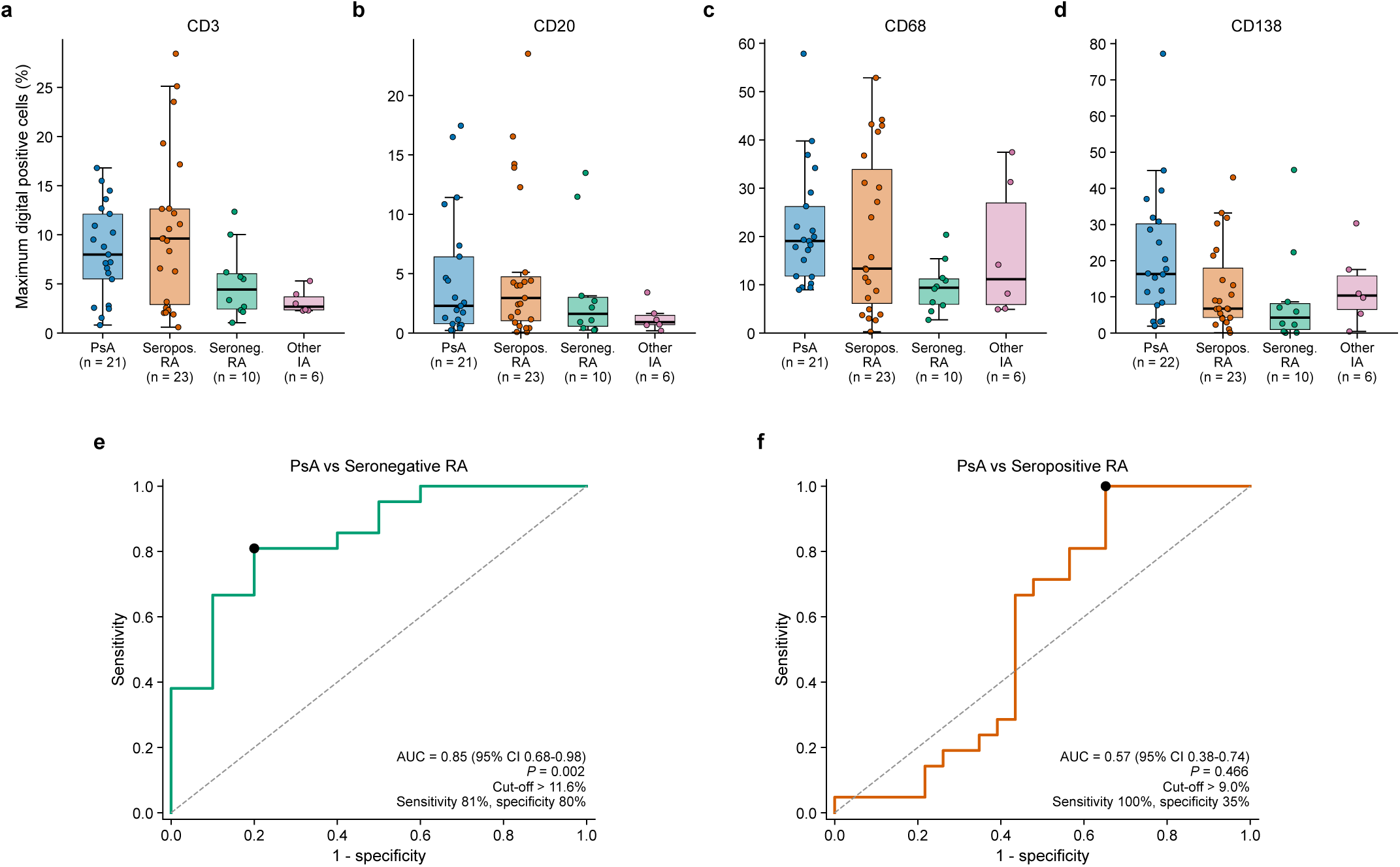
Digital pathology scores for the four conventional pathotype immunohistochemistry markers by diagnosis revealing a CD68 marker signature differentiating PsA from seronegative RA. (a–d) Boxplots of individual immunohistochemical marker scores based on % positive cells by diagnosis for (a) CD3, (b) CD20, (c) CD68 and (d) CD138. (e, f) ROC curve post-hoc analysis with CD68 positivity as the classifier to differentiate PsA from (e) seronegative or (f) seropositive RA.

### Vascularity and fibrosis are comparable across inflammatory arthritides

Vascularity and fibrosis were quantified using CD31 vascular area fraction and proportional fibrosis area (picrosirius red with fast green staining), respectively. Comparison of these parameters by diagnosis revealed similar levels of vascularity and stromal fibrosis across the diseases, with no significant divergence noted between PsA and seropositive RA (Figures 3b–e, Supplementary Table 3).

**Figure 3.**
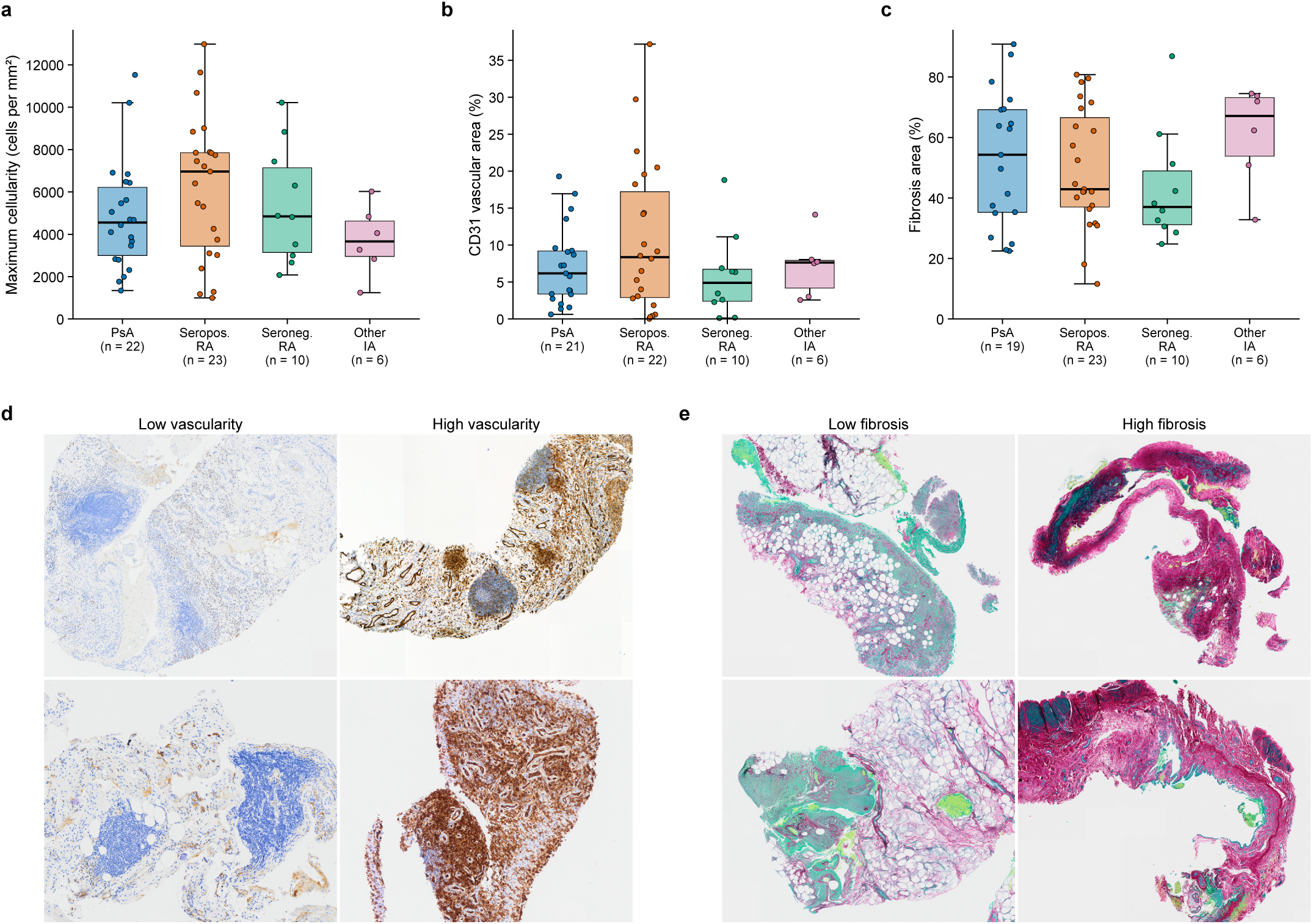
Assessment of digital pathology scores from H&E, CD31 and Picrosirius Red staining reveals similar levels of cellularity, vascularity and fibrosis between diseases. (a) Boxplots of cellularity by diagnosis. (b, c) Boxplots of (b) vascularity as determined by vascular area fraction (area of CD31 staining / total tissue area) and (c) fibrosis (area of collagen staining in red / total tissue area) by diagnosis. (d) Representative synovial tissue images from patient samples with low and high vascularity based on CD31 staining. (e) Representative synovial tissue images from patient samples with low and high fibrosis based on picrosirius red with fast green staining.

### The pauci-immune pathotype is predominantly identified in patients receiving treatment

Patients were recruited across the clinical spectrum of disease: 31% were treatment-naïve (n=19/61), 31% were receiving conventional synthetic DMARDs (csDMARDs; n=19/61), and 38% were receiving biologic or targeted synthetic DMARDs (b/tsDMARDs; n=23/61) (Table 1, Figure 4a). Integrating treatment history with pathotype assignments revealed that the pauci-immune pathotype was pre-dominantly identified in treatment-experienced patients (those receiving csDMARDs or b/tsDMARDs; n=7/8), with one treatment-naïve patient assigned this pathotype (Figures 4b–f), although the overall distribution of pathotypes did not differ significantly by treatment (chi-squared p=0.690). However, comparison of individual continuous histological markers across treatment groups found no significant differences between patients receiving b/tsDMARDs, csDMARDs, or those who were treatment-naïve (Supplementary Figures 8a and 8b).

**Figure 4.**
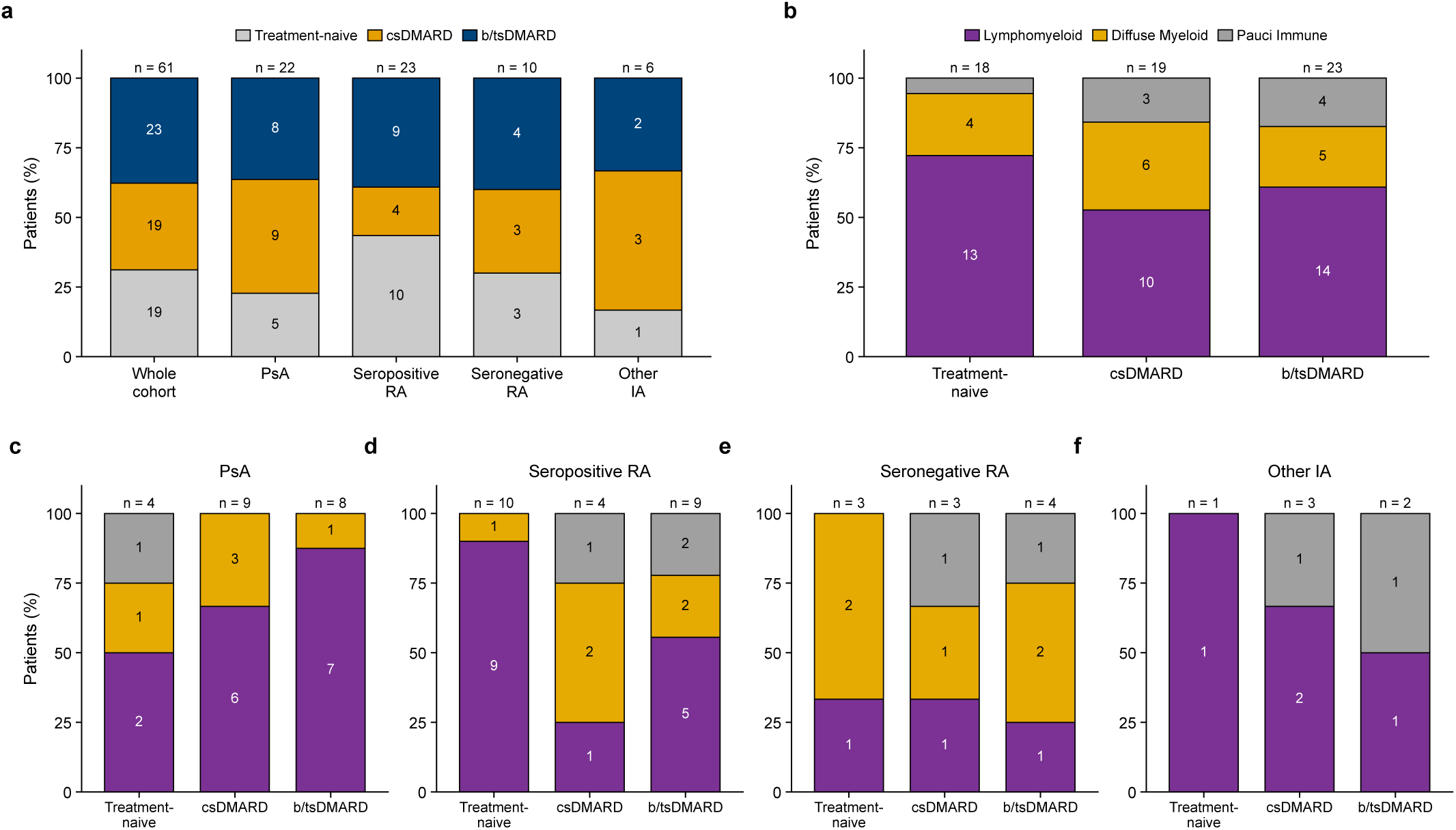
Assessment of RA synovial pathotypes stratified by treatment and diagnosis reveals a numerically higher proportion of patients with a pauci immune pathotype amongst those receiving treatment compared to treatment naïve patients. (a) Overview of the patient cohort by treatment regimen overall and by diagnosis. (b) Proportions of RA synovial pathotypes observed by treatment regimen. (c–f) Proportions of RA synovial pathotypes observed by treatment regimen in (c) PsA, (d) seropositive RA, (e) seronegative RA and (f) other inflammatory arthritis.

### Synovial pathotypes and histological profiles are independent of disease duration

Neither individual histological profiles nor assigned synovial pathotypes demonstrated a significant correlation with disease duration. This lack of temporal association suggests that these pathotypes represent distinct, intrinsic biological disease states rather than sequential stages of disease progression (Supplementary Figures 9a–h).

### A subset of PsA patients shares the inflammatory synovial profile of seropositive RA

To investigate histological overlap between clinical diagnoses, unsupervised clustering of the entire cohort (n=60) was performed utilising all seven histological markers. Cluster number was evaluated via the elbow method and silhouette analysis, and three distinct phenotypic groups were defined. Cluster 1 was characterised by high cellularity, vascularity, and CD3/CD20/CD68 expression; Cluster 2 by high vascularity alongside prominent CD138 and CD68 expression; and Cluster 3 by profound fibrosis. Evaluating the diagnostic distribution across these profiles revealed that most seropositive RA patients mapped to the highly inflammatory Cluster 1, alongside 33% (n=7/21) of the PsA cohort. Conversely, PsA patients comprised the largest proportion of Cluster 2 (50%, n=5/10), whereas Cluster 3 contained similar proportions of PsA, seropositive and seronegative RA patients. Together, these data demonstrate that a subset of PsA patients share a highly inflamed, vascular, and lymphocyte/myeloidrich synovial phenotype with seropositive RA, whereas a distinct subset of PsA patients have synovial inflammation characterised by high CD138 expression (Supplementary Figures 10a–c).

To further dissect this inflammatory overlap, a secondary unsupervised clustering analysis was performed exclusively on patients categorised with a classic lymphomyeloid pathotype (n=37). This sub-clustering yielded two distinct groups. Cluster 1 exhibited traditional inflammatory hallmarks (higher CD3, CD20, CD68, vascularity, and overall cellularity), whereas Cluster 2 was distinguished by disproportionately high CD138 expression and fibrosis. Diagnostic stratification of this sub-cohort revealed a clear divergence: 73% (n=11/15) of the lymphomyeloid seropositive RA patients localised to Cluster 1, accompanied by 40% (n=6/15) of the lymphomyeloid PsA patients. By contrast, Cluster 2 was predominantly composed of PsA patients (50%, n=9/18) and only 22% (n=4/18) seropositive RA patients. These findings indicate that while roughly half of lymphomyeloid PsA patients exhibit an inflammatory signature indistinguishable from seropositive RA, the remainder represent a distinct pathological subgroup driven by CD138^+^ plasma cell infiltration and stromal remodelling (Supplementary Figures 11a–c).

### Unsupervised clustering defines three novel, clinically distinct synovial pathotypes in PsA

To deconstruct the heterogeneity of synovial inflammation specifically within the PsA cohort (n=22), unsupervised hierarchical clustering was performed utilising all seven histological markers. This yielded three distinct phenotypic clusters, which were assigned descriptive labels based on their defining histological architecture. Cluster 1, driven by high CD68 expression and dense overall cellularity, was designated the ‘Dense Myeloid’ pathotype. Cluster 2, defined by profound fibrosis and a relative paucity of immune infiltration (low CD3, CD20, CD68, and CD138), was classified as the ‘Fibro-Stromal’ pathotype. Finally, Cluster 3, distinguished by pronounced CD138^+^ plasma cell infiltration and increased vascularity, was named the ‘Angio-Lymphoid’ pathotype (Figures 5a and 5c, Supplementary Figures 12a–c). Evaluation of baseline medication exposure revealed that treatment modalities were broadly distributed across all three pathotypes, suggesting these histological profiles represent intrinsic disease states rather than artifacts of specific therapeutic regimens (Figure 5a, table in Supplementary Figure 12).

**Figure 5.**
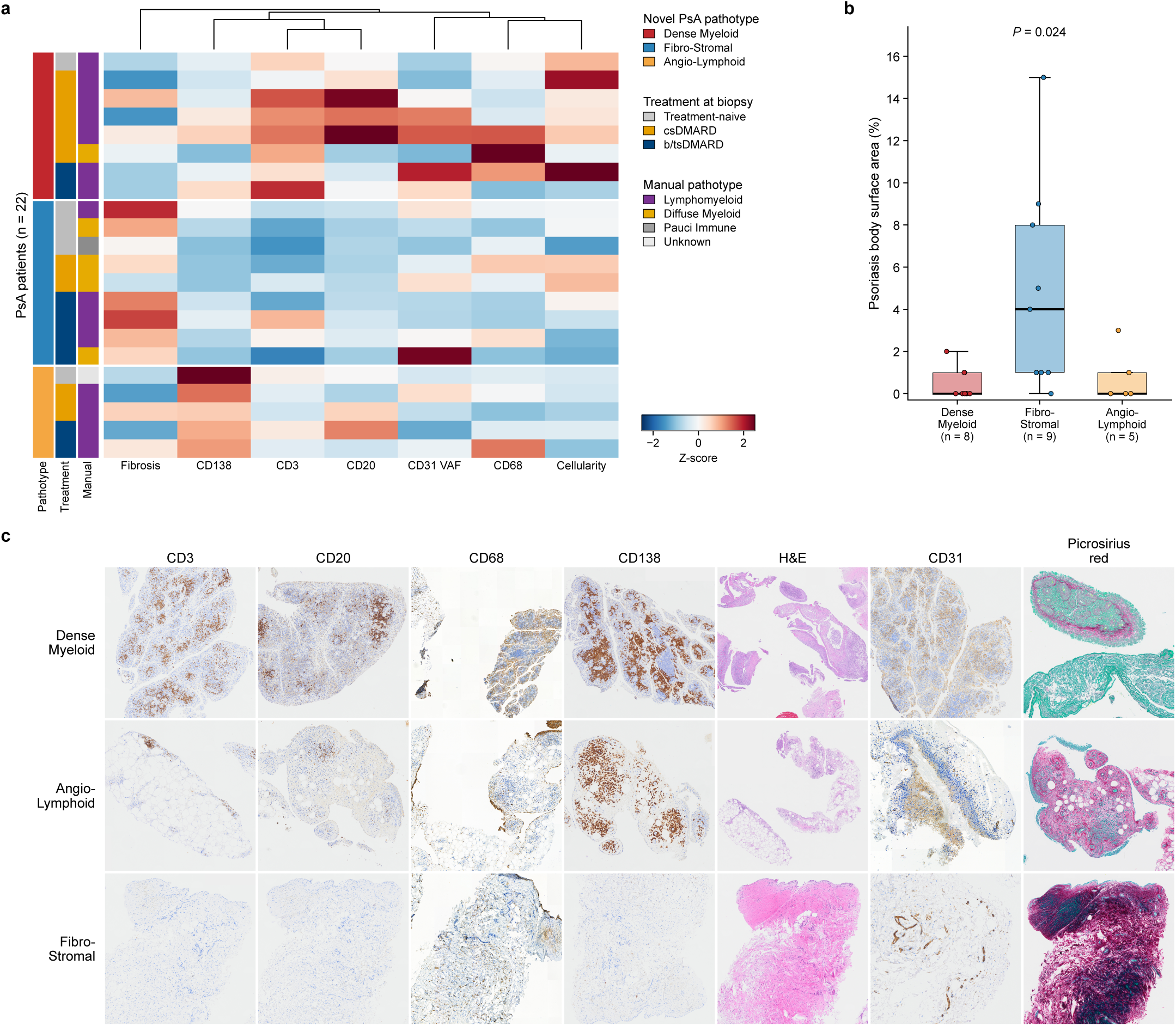
Machine learning approaches reveal novel PsA pathotypes. (a) Unsupervised hierarchical clustering heatmap derived from synovial histological markers across the PsA cohort (n = 22), establishing three distinct pathotypes: Dense Myeloid, Angio-Lymphoid, and Fibro-Stromal. (b) Psoriasis body surface area by novel PsA pathotype; P is the unadjusted Kruskal-Wallis P value. (c) Representative histological images illustrating the unique structural, vascular, and inflammatory features characterising each of the three novel pathotypes.

Integrating these novel pathotypes with clinical measures of disease activity revealed a striking divergence between joint and skin phenotypes. Patients exhibiting the Fibro-Stromal pathotype demon-strated a significantly higher psoriasis body surface area (BSA) compared to the other groups (Figure 5b, Supplementary Figure 13e, Supplementary Table 4). Evaluating the individual histological markers as continuous variables further reinforced this relationship. CD138 expression correlated positively with both patient and physician global assessments of disease severity (Supplementary Figures 14a and 14b); however, both CD138 expression and overall synovial cellularity correlated negatively with cutaneous psoriasis severity (BSA) (Supplementary Figures 14c and 14e). Conversely, the proportional area of synovial fibrosis demonstrated a significant positive correlation with BSA (Supplementary Figure 14d). Together, these findings indicate that while highly cellular, plasma cell-rich synovial inflammation drives systemic and joint-associated disease activity, severe cutaneous psoriasis is associated with a fibrotic, pauci-immune synovial landscape (Supplementary Figures 13a–g and 14a–e, Supplementary Tables 4 and 5).

### A clinically translatable decision tree algorithm for routine PsA synovial pathotyping

To facilitate the integration of these novel PsA synovial pathotypes into routine clinical histopathology workflows, a diagnostic decision tree algorithm was developed (Figure 6a). This algorithm combines standard H&E cellularity assessment with a focused four-marker panel (CD31, CD68, CD138, and picrosirius red with fast green) utilising standard semi-quantitative scoring (0–3). Marker selection was driven by a combination of capacity for cluster discrimination and practical alignment with existing RA pathotyping protocols, which traditionally rely on a four-marker plus H&E framework.

**Figure 6.**
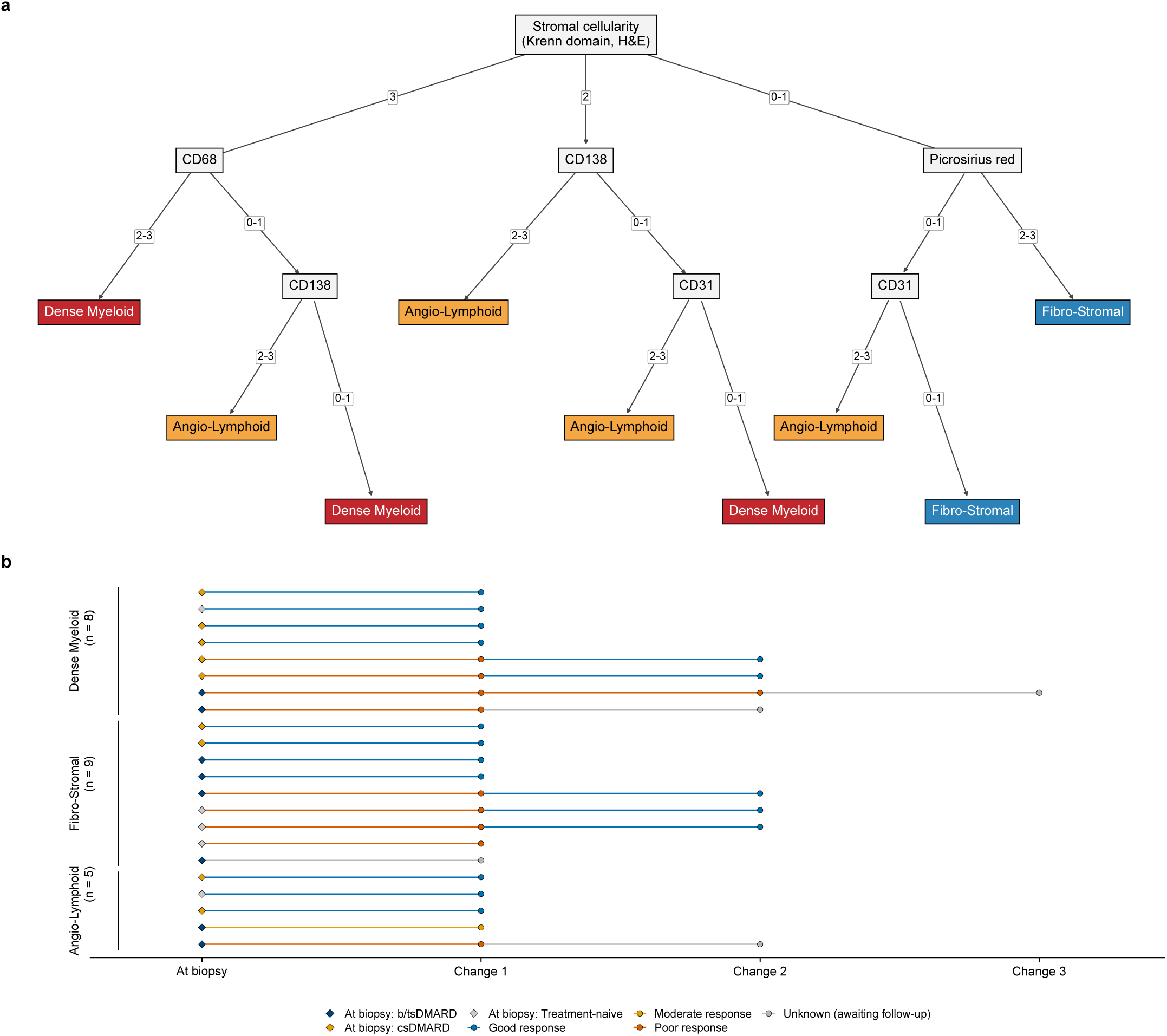
Novel PsA pathotypes are adaptable into a focused 4-marker panel for clinical applications, and treatment response outcomes by novel pathotype. (a) A clinically translatable decision tree algorithm combining standard H&E assessment with a focussed 4-marker panel (CD31, CD68, CD138, and Picrosirius Red) to facilitate pathotype assignment in routine clinical histopathological workflows. (b) Swimmer plot of longitudinal treatment response outcomes stratified by novel PsA pathotype for the PsA cohort.

Specifically, because unsupervised clustering revealed that plasma cell (CD138) and vascular (CD31) expression were the primary drivers differentiating the Dense Myeloid and Angio-Lymphoid pathotypes, CD138 was incorporated into the final algorithm in lieu of the traditional B-cell marker CD20. Picrosirius red was introduced to reliably isolate the highly fibrotic architecture of the Fibro-Stromal pathotype. Finally, as the sole myeloid marker in the panel, CD68 was retained to ensure robust, high-confidence identification of the Dense Myeloid phenotype (Figure 6a).

### Longitudinal treatment outcomes and pathotype-specific therapeutic responses in PsA

Across the entire biopsy cohort (n=61), the median follow-up duration from the time of biopsy was 2.11 years (interquartile range [IQR] 1.21–3.05), with treatment trajectories for the entire cohort shown in Supplementary Figure 15. Stratification by diagnosis revealed a slightly shorter follow-up duration for the PsA cohort (median 1.44 years, IQR 1.02–2.33) compared to the seropositive RA cohort (median 2.45 years, IQR 1.69–4.14) (Supplementary Figure 16c, Supplementary Table 6). Following biopsy, all 22 PsA patients underwent a change in their therapeutic regimen. The most common primary interventions were methotrexate (41%, n=9/22) and adalimumab (36%, n=8/22), followed by secukinumab (n=2/22), abatacept (n=1/22), tacrolimus (n=1/22), and infliximab (n=1/22) (Supplementary Figure 16b, Supplementary Table 7). Evaluating longitudinal clinical efficacy revealed that 55% of the PsA cohort (n=12/22) achieved a good or moderate response and maintained their initial post-biopsy therapy at their most recent follow-up. Conversely, 41% (n=9/22) demonstrated a poor response of which 89% (n=8/9) underwent subsequent switch in treatment regimen. One patient did not have longitudinal treatment outcomes available as they are awaiting clinical follow-up assessment. (Figure 6b, Supplementary Figure 16a, Supplementary Table 8)

We next investigated whether the novel baseline synovial pathotypes were associated with this subsequent therapeutic failure. A slightly lower proportion of patients within the Fibro-Stromal pathotype exhibited a good or moderate longitudinal treatment response (44.4%, n=4/9) compared to those in the Dense Myeloid (50.0%, n=4/8) and Angio-Lymphoid (80.0%, n=4/5) cohorts. Although this trend aligns with the highly fibrotic, treatment-resistant phenotypes often observed in pauci-immune joint disease, the differences did not reach statistical significance, likely due to the limited statistical power inherent in the cohort size (Figure 6b, Supplementary Table 8).

### Novel synovial pathotypes are independent of concurrent radiographic osteoarthritis

Synovial inflammation is increasingly recognised as a feature of primary osteoarthritis (OA).^40, 41^ Given that PsA frequently co-occurs with OA, and because our PsA cohort was biopsied exclusively at the knee (Supplementary Figures 17e and 17f), which is a weight-bearing joint highly susceptible to degenerative change, we evaluated concurrent radiographic OA severity at the time of biopsy to exclude it as a confounding driver of our clustering. Radiographs were available for 17 of the 22 PsA patients. Of these, 29% exhibited no radiographic OA (n=5/17), with the remainder demonstrating mild OA (47%, n=8/17) or moderate OA (24%, n=4/17) (Supplementary Figure 17a). Notably, no PsA patients in our cohort exhibited severe radiographic OA. While stratification of OA severity confirmed expected physiological trends, with moderate degenerative changes occurring more frequently in older patients and those with longer disease durations, there was no association between OA severity and the assigned PsA pathotype. Crucially, radiographic OA was distributed proportionally across all three clusters, suggesting that these distinct synovial signatures, including the highly fibrotic Fibro-Stromal phenotype, represent primary psoriatic disease mechanisms rather than secondary degenerative artifacts (Supplementary Figures 17b–d).

## Discussion

We present the first computationally derived, PsA-specific synovial pathotypes. By leveraging objective digital pathology to move beyond traditional RA-centric models, we defined three distinct pathotypes to characterise synovial inflammation in PsA: Dense Myeloid, Fibro-Stromal, and Angio-Lymphoid. While a subset of PsA patients shares an inflammatory signature with seropositive RA, applying RA pathotypes to the broader PsA cohort fails to capture its unique fibrotic and plasma-cell-driven architecture. To bridge computational discovery and clinical utility, we translated these findings into a pragmatic, four-marker diagnostic decision tree to facilitate future tissue-driven studies in PsA.

A key biological insight is the prominent role of CD138^+^ plasma cells in the inflammatory PsA synovial microenvironment. Unlike the CD20^+^ B cell-dominated RA lympho-myeloid pathotype, sub-clustering revealed a substantial subset of inflamed PsA characterised by disproportionate CD138^+^ plasma cell infiltration and hypervascularity. This distinct Angio-Lymphoid signature justified substituting CD20 with CD138 in our PsA-specific algorithm. This plasma cell enrichment aligns with previously published evidence of profound B-cell maturation in psoriatic disease,^8, 34, 42, 43^ suggesting intra-articular terminal B-cell differentiation may be a defining feature of severe PsA synovitis.

Clinically, these pathotypes revealed a striking divergence between joint and skin phenotypes. Synovial immune infiltration correlated inversely with cutaneous severity: patients with the pauci-immune Fibro-Stromal pathotype exhibited higher psoriasis BSA, whereas dense cellularity and CD138 expression correlated with lower BSA. This divergence aligns with real-world observations where approximately 34% of PsA patients exhibit severe joint but mild skin involvement,^44^ suggesting a tissue-specific compartmentalisation of the immune response. Importantly, systematic radiographic evaluation confirmed the Fibro-Stromal signature is an intrinsic PsA feature, as concurrent knee osteoarthritis severity was distributed evenly across all pathotypes and decoupled from fibrosis.

Longitudinally, these pathotypes demonstrate potential to guide rational drug selection. Following biopsy, patients predominantly initiated methotrexate or TNF inhibitors. Fibro-Stromal patients trended towards higher subsequent treatment failure, where only 44% had a good or moderate response to treatment compared to 50% of the PsA patients with a Dense Myeloid pathotype and 80% of the PsA patients with a Angio-Lymphoid pathotype. Although underpowered for statistical significance, this trend provides a compelling biological rationale: standard therapies may be sub-optimal for this skin-predominant, pauci-immune, and fibrotic phenotype. Because IL-17 and IL-23 inhibitors demonstrate superior efficacy for skin clearance compared to TNF inhibition,^45, 46^ the refractory nature of the Fibro-Stromal pathotype may reflect a mechanistic mismatch. This highlights the need for a prospective study to explore the actionable opportunity to stratify these patients towards early IL-17/IL-23 inhibitor interventions, alongside an unmet need for novel therapeutics targeting pathogenic synovial fibroblasts.

Study limitations include the modest PsA cohort size (n=22), which constrained the statistical power of longitudinal analyses and necessitated grouping diverse drug classes. Validation in larger, prospectively treated cohorts is required to confirm predictive values for specific biologics. Additionally, while exclusively sampling the knee standardised the anatomical site and we rigorously excluded OA as a confounder, it remains unclear whether these pathotypes are uniformly represented in smaller joints. Finally, our multi-modal histological framework is restricted to predefined markers. Future studies leveraging single-cell and spatial transcriptomics^30, 31, 47–52^ will be crucial. Just as single-cell transcriptomics in RA discovered highly granular Cell-Type Abundance Phenotypes (CTAPs),^53^ multi-omic technologies will inevitably refine these PsA pathotypes.

Ultimately, establishing data-driven, PsA-specific synovial pathotypes marks a pivotal step toward precision medicine. By moving away from RA-centric models, we provide a framework that accurately reflects the distinct plasma cell and fibrotic architecture of PsA. Our pragmatic diagnostic decision tree, if independently replicated, is designed for immediate integration into routine clinical histopathology without requiring advanced transcriptomics. The true potential of this framework lies in its application in biomarker-stratified clinical trials. Aligning the expanding PsA therapeutic armamentarium with intrinsic synovial biology is essential to replace empirical trial-and-error medicine with targeted, tissue-driven care.

## Contributors

All authors contributed to the conceptualisation of the study. RMH designed the methodology, conducted the laboratory experiments, curated the data, generated the figures, and wrote the original draft of the manuscript. MC, SLS, YT, and AW assisted with the laboratory experiments and data generation. Clinical investigation, including patient recruitment and the performance of ultrasoundguided synovial biopsies, was conducted by RMH, YT, RS, and PH. LMM, MS, and PM contributed to the formal data analysis. GO, SV, MB, AB, PM, and PH provided critical oversight, expertise, protocols, samples, research funding, and supervised the research. All authors contributed to the review and editing of the revised manuscript. All authors had full access to all the data in the study and share final responsibility for the decision to submit the manuscript for publication.

## Declaration of generative AI and AI-assisted technologies in the manuscript preparation process

During the preparation of this work, the authors used Google Gemini and Claude (Anthropic) for language editing, grammatical correction, and stylistic refinement of selected sections. Claude was also used to assist with reviewing and correcting analysis code, running the statistical analyses, generating figures and tables, and updating the reported results. The authors designed the study, reviewed and verified all outputs, drew the scientific conclusions, and take full responsibility for the content and academic integrity of the final publication.

## Data sharing

Anonymised individual participant data will be available from the corresponding author upon reasonable request following publication, subject to approval by the relevant national and institutional authorities and applicable data protection regulations.

## Acknowledgements

The authors would like to express their sincere gratitude to all the patients who generously donated tissue and participated in this research, as well as the dedicated clinicians involved in their care. We also thank the Kellgren Centre for Rheumatology research team, including the research nurses and study coordinators, alongside the university study coordinators, for their invaluable logistical and administrative support. We extend our thanks to VOCAL for their vital guidance and support regarding patient and public involvement. We gratefully acknowledge the technical expertise and assistance provided by The University of Manchester Histology and Bioimaging Core Facility (Peter March, Abigail Byford, Nicole Campbell, Karolina Krakowiak, and Steven Marsden), as well as the essential contributions of our research technicians (Eryn Doe, Justin Engelbert, and Amirah Binti Mohamed Ariff).

The authors acknowledge the support received from Versus Arthritis (21754, 21818) and the National Institute for Health and Care Research (NIHR) Manchester Biomedical Research Centre (NIHR203308). The views expressed are those of the authors and not necessarily those of the NIHR. RMH is supported by a Wellcome Trust 4ward North Clinical PhD Fellowship (323724/Z/24/Z), a BritPACT pilot research grant, and a National Psoriasis Foundation Psoriatic Disease Research Fellowship. LMM is funded by the Medical Research Council (MRC) through the MRC Doctoral Training Partnership (MR/W007428/1).

