## Supplementary Materials for "Redefining Synovial Pathotypes in Psoriatic Arthritis through Multi-Modal Histology and Machine Learning (The PSYNOVIUM Study)"

Supplementary Figure 9. Disease duration shows no correlation with histological markers or pathotypes 16

Supplementary Figure 10. Unsupervised clustering of the entire cohort does not show clear clustering by diagnosis or RA pathotype, suggesting similarities between PsA and RA synovial inflammation 17

**References** **29**

### Supplementary methods

*Patient Recruitment and Clinical Data Collection*

This cross-sectional, multi-modal study was designed to characterise the synovial microenvironment in Psoriatic Arthritis (PsA) compared to seropositive Rheumatoid Arthritis (RA) and other forms of inflammatory arthritis. Patients were recruited from The Kellgren Centre for Rheumatology at the Manchester Royal Infirmary, Manchester University NHS Foundation Trust, which operates as the sole tertiary synovial biopsy referral centre in the North West of England. Biological samples were acquired from patients enrolled across several observational cohorts, including the National Repository study (REC No: 99/8/84 IRAS 32231), the PRECISE-RA study (REC No: 20/SS/0090 IRAS 281253), and the SYNPATIA study (REC No: 16/NW/0571 IRAS 186499). Ethical approval was granted by the respective research ethics committees. All participants provided informed written consent prior to inclusion.

Adult patients presenting with ultrasound-detected inflammation (grey-scale $\geq$2 and power Doppler $\geq$1) in a joint amenable to needle core biopsy (knee, wrist, elbow, or ankle) were eligible for inclusion. Diagnostic inclusion for analysis required PsA patients to fulfil the CASPAR classification criteria,[1] while seropositive RA patients were required to fulfil the 2010 ACR/EULAR classification criteria[2] with a strictly defined positive anti-citrullinated protein antibody (ACPA) titre, independent of rheumatoid factor status. Seronegative RA was defined as RA with a negative ACPA titre, independent of rheumatoid factor status. Comprehensive baseline clinical metrics including psoriasis body surface area (BSA), joint counts, and acute phase reactants were recorded at the time of biopsy. Furthermore, to evaluate potential structural confounding, concurrent radiographic osteoarthritis (OA) severity at the biopsied joint was assessed from pre-biopsy imaging and graded as none, mild, moderate, or severe based on a radiologist report. Longitudinal therapeutic efficacy was evaluated using UK NICE guidelines, defined by the Psoriatic Arthritis Response Criteria (PsARC) for PsA patients[3] and EULAR response criteria (derived from DAS28) for RA patients.[4]

*Ultrasound-Guided Synovial Biopsy*

Synovial biopsies were performed in accordance with established local protocols. The target joint was initially assessed via ultrasound to confirm adequate synovial hypertrophy ($\geq$1 cm) and the presence of an effusion. Following the establishment of a sterile field, local anaesthesia was administered using ethyl chloride spray followed by subcutaneous and intra-articular infiltration of 1% lidocaine. Synovial fluid was aspirated prior to tissue collection. A trocar was subsequently introduced into the joint space, and the joint was distended with 5–10 mL of 1% lidocaine to optimize tissue accessibility. A 14-gauge core biopsy needle was advanced through the trocar to systematically harvest synovial biopsy cores (1.6 mm diameter, up to 10 mm length). To minimize patient discomfort and perioperative complications, the procedure was terminated after the collection of a maximum of 40 cores or after 30 minutes. Extracted cores were immediately transferred into 10% neutral buffered formalin for fixation, or into chilled PBS/complete cell media (RPMI) on ice and transported directly to the university laboratory for processing within one hour of collection.

*Tissue processing and embedding*

Synovial tissue cores designated for histological analysis were fixed in 10% neutral buffered formalin for a minimum of 24 hours to preserve cellular architecture before being stored in 70% ethanol until tissue processing. Fixed tissues were transferred to the University of Manchester Histology Core Facility for automated processing using a Leica ASP300 S Fully Enclosed Tissue Processor (Leica Biosystems). The overnight processing protocol comprised:

- Dehydration: Sequential immersion in graded ethanol baths (70%, 90%, and three exchanges of 100%) to completely displace aqueous fluid.
- Clearing: Three sequential immersions in xylene to displace ethanol and prepare the tissue matrix for wax infiltration.
- Infiltration: Impregnation with molten paraffin wax at 60°C under negative pressure to ensure complete cellular stabilisation.

Processed samples were manually embedded into paraffin blocks using a Leica EG1150 Embedding Centre. Biopsy cores were oriented parallel to the cutting face to ensure maximum cross-sectional tissue surface area during microtomy.

*FFPE Tissue Storage and Slide Preparation*

Formalin-fixed paraffin-embedded (FFPE) blocks were catalogued and maintained at 4°C. Blocks were faced to expose the full tissue cross-section using a Leica HistoCore MULTICUT rotary microtome (Leica Biosystems), after which serial sections were cut at a thickness of 5 µm. Tissue ribbons were floated on a room-temperature water bath for initial manipulation, followed by transfer to a 40°C heated water bath for expansion. Sections free of structural artifacts were mounted onto SuperFrost adhesive glass slides (Epredia), with 2–4 consecutive tissue sections placed per slide to maximize tissue area. A minimum of seven sequential slides were generated per block. Slides were labelled via an automated system and dried at room temperature for 24 hours to ensure robust tissue adhesion.

*Multi-Modal Staining Strategy*

A multi-modal histological approach was deployed across sequential tissue sections to independently capture the three primary domains of synovial pathology: general inflammation (Haematoxylin & Eosin), structural stromal fibrosis (Picrosirius Red with Fast Green), and specific immune/vascular phenotyping (Immunohistochemistry). Due to finite tissue volumes in standard needle core biopsies, a strict staining hierarchy was implemented for blocks with low residual capacity: H&E was prioritised first, followed by Picrosirius Red, and subsequently the IHC panel ordered by biological priority (CD20 → CD68 → CD138 → CD3 → CD31).

*Histochemical Staining (H&E)*

Automated H&E staining was executed using a Leica ST5010 Autostainer XL workstation integrated with a Leica CV5030 Glass Coverslipper. Sections were deparaffinised in xylene and rehydrated through a graded ethanol series to distilled water. Nuclear staining was achieved by immersion in Haematoxylin (Gill II) for 5–7 minutes, followed by differentiation in 1% acid alcohol and bluing in Scott's Tap Water Substitute. Cytoplasm was counterstained with Eosin Y (alcoholic) for 1–2 minutes. Sections were dehydrated, cleared in xylene, and coverslipped using CV Mount medium.

*Picrosirius Red Staining for Fibrosis Quantification*

To quantify synovial extracellular matrix deposition, Picrosirius Red staining with a Fast Green counterstain was utilised. Sirius Red (in picric acid) specifically binds to collagenous networks (primarily types I and III), yielding a bright red stain, while Fast Green provides high-contrast background staining of non-collagenous proteins and cytoplasm. This stark optical contrast facilitates highly accurate digital segmentation of the fibrotic area fraction compared to standard H&E staining.

*Immunohistochemistry (IHC)*

IHC was performed on the automated Leica BOND Rx platform (Leica Biosystems). The antibody panel (Supplementary Methods Table 1) was designed to capture T cells (CD3), B cells (CD20), macrophages (CD68), plasma cells (CD138), and endothelial cells (CD31), expanding upon established RA pathotyping criteria to capture the prominent vascularity and plasma cell infiltration hypothesized to define PsA synovitis.

**Supplementary Methods Table 1. Immunohistochemistry antibody panel**

| Target | Clone | Vendor | Dilution | HIER Buffer | Reference |
|---|---|---|---|---|---|
| CD3 | 2GV6 | Ventana/Roche | RTU | ER1 (Citrate) | Rao et al. 2017[5] |
| CD20 | L26 | Ventana/Roche | RTU | ER1 (Citrate) | Rivellese et al. 2022[6] |
| CD68 | PG-M1 | Agilent/Dako | 1:100 | ER2 (EDTA) | Rivellese et al. 2022[6] |
| CD138 | MI15 | Agilent/Dako | 1:100 | ER2 (EDTA) | Rivellese et al. 2022[6] |
| CD31 | JC70A | Agilent/Dako | 1:100 | ER2 (EDTA) | Wei et al. 2020[7] |

*Note: Ready-to-Use (RTU) Ventana CONFIRM dispensers natively designed for the Roche BenchMark platform were manually decanted into Leica-compatible 7mL Open Containers. This platform-agnostic approach preserved validated clinical clones while leveraging the fluidics and heat-retrieval flexibility of the BOND RX platform.*

Slides were dewaxed utilising Bond Dewax Solution (Leica Biosystems). Heat-induced epitope retrieval (HIER) was executed using Epitope Retrieval Solution 1 (citrate, pH 6.0) or Solution 2 (EDTA, pH 9.0) as per optimised clone requirements. Detection was achieved via the Bond Polymer Refine Detection system, utilising a horseradish peroxidase (HRP) linker and 3,3'-Diaminobenzidine (DAB) chromogen, followed by a haematoxylin nuclear counterstain.

*Whole-Slide Image Acquisition and Digital Pathology*

Whole slide images (WSIs) were acquired at 20× magnification using either a 3DHistech Panoramic 250 Flash

III (0.24 µm/pixel) or an Olympus SLIDEVIEW VS200 (0.274 µm/pixel) slide scanner. Digital image analysis was conducted blindly using QuPath software (v0.6.0).

*Cellularity and Krenn Synovitis Scoring*

Tissue images were digitally annotated to exclude artifacts and define the evaluable surface area. QuPath's watershed cell detection algorithm was deployed on H&E WSIs, utilising optical density thresholding in the haematoxylin channel to identify distinct nuclei. Total digital cellularity was calculated as the absolute nuclear count normalised to the evaluable tissue area (cells/mm$^2$). Concurrently, H&E sections were subjected to standardised Krenn synovitis scoring (range 0–9), evaluating lining layer hyperplasia, stromal cellularity, and inflammatory infiltrate (Supplementary Methods Table 2). Grading was performed blindly by three independent observers, with discrepancies resolved via joint adjudication.

**Supplementary Methods Table 2. Criteria for the Krenn synovitis score**

| Score | Lining Hyperplasia | Stromal Cellularity | Inflammatory Infiltrate |
|---|---|---|---|
| 0 | 1–2 cells | Normal cellularity; sparse resident cells with abundant extracellular matrix. | No inflammatory infiltrate or widely scattered single immune cells. |
| 1 | 3–4 cells | Mild activation; noticeable increase in resident cells but stroma remains loose. | Mild infiltration; scattered lymphocytes or small focal perivascular aggregates. |
| 2 | 5–9 cells | Moderate activation; cellular crowding with a loose, interconnected network. | Moderate infiltration; numerous small-to-medium lymphoid follicles or diffuse spread. |
| 3 | >10 cells | Severe activation; densely packed resident cells forming solid sheets (pannus). | Severe infiltration; massive lymphoid follicles or dense, architecture-obscuring diffuse infiltrates. |

*Machine Learning-Driven Pixel Classification (Fibrosis and Vascularity)*

To quantify structural tissue remodelling, supervised machine learning was utilised. High-resolution Random Trees pixel classifiers were trained within QuPath on a representative 10% subset of WSIs. For Picrosirius Red sections, the classifier was trained utilising image features to segment pixels into Collagen, Tissue, Blood, and Background classes. Proportional fibrotic burden (fibrosis %) was defined as the total Collagen area divided by the total evaluable Tissue area. An analogous Random Trees classifier was trained on CD31 IHC sections to isolate DAB-positive endothelial structures and extract the vascular area fraction (VAF %).

*Immunohistochemical Cell Detection, Semi-Quantitative Grading, and Pathotype Assignment*

For sections stained with CD3, CD20, CD68, and CD138, digital quantification was executed using QuPath's positive cell detection tool to derive absolute positive cell counts and proportional positivity rates. In parallel with this digital analysis, traditional semi-quantitative grading (0–3) was performed for all lineage markers and structural stains to establish a clinical ground truth. To ensure rigorous inter-observer reliability, grading was conducted blindly by three independent observers. Any discrepancies were subsequently resolved via joint adjudication to achieve a unanimous consensus score.

Specific grading criteria were defined for the distinct histological domains:

- Immune Infiltration (CD3, CD20, CD68, CD138): The degree of synovial infiltration within the sublining and lining layers was graded as: Grade 0 (absent or rare scattered positive cells); Grade 1 (mild, scattered infiltration or occasional loose clusters); Grade 2 (moderate, multifocal aggregates or distinct perivascular cuffing); and Grade 3 (severe, dense confluent sheets or large, highly organised lymphoid aggregates).
- Synovial Vascularity (CD31): Endothelial density and prominence within the sublining layer were graded as: Grade 0 (minimal staining with rare distinct vessels); Grade 1 (occasional scattered vessels with minor background staining); Grade 2 (moderate, clear vascularity with several distinct vessels); and Grade 3 (prominent staining with numerous vessels distributed densely throughout the sublining tissue, indicative of pronounced, tortuous angiogenesis).
- Synovial Fibrosis (Picrosirius Red): Collagen deposition on Fast Green-counterstained sections was graded as: Grade 0 (no red collagen staining); Grade 1 (mild, focal fibrosis involving <25% of the evaluable tissue area); Grade 2 (moderate, clear fibrosis affecting approximately 50% of the synovial tissue); and Grade 3 (extensive, significant fibrosis where the majority of the tissue architecture [>50%] was densely packed with intensely stained red collagen structures).

Following manual evaluation, the consensus semi-quantitative grades for the four standard lineage markers (CD3, CD20, CD68, CD138) were utilised to assign established synovial pathotypes, adhering to the classification framework described by Humby et al.[8] To bridge traditional histopathology with computational image analysis, these manual assignments were directly correlated with the continuous variables generated by the digital IHC pipeline.

*Data Aggregation and Feature Matrix Construction*

To construct a comprehensive, multi-dimensional profile of the synovial microenvironment, both quantitative digital pathology metrics and manual semi-quantitative grades were aggregated at the patient level. The final feature matrix comprised a dual-modality dataset. The continuous digital variables included overall tissue cellularity (cells/mm$^2$), vascular area fraction (%), proportional fibrotic burden (%), and the positive detection rates for specific lineage markers (CD3, CD20, CD68, and CD138). These computational metrics were integrated alongside their corresponding manually assigned semi-quantitative scores (grades 0–3) for vascularity, fibrosis, and individual immunohistochemical infiltration. To ensure the downstream analysis accurately captured the peak lesional severity of the inflamed joint, the maximum regional value, whether the continuous digital metric or the discrete manual grade, across all evaluable biopsy blocks was selected for each respective feature per patient.

*Missing Data Imputation and Feature Scaling*

Patients missing more than two of the seven requisite histological markers were excluded from the downstream analysis to preserve dataset integrity, with one exception: one PsA patient with five of the seven markers missing was excluded from whole-cohort clustering but retained in the PsA-only clustering. For tissues with missing variables, missing data points were imputed utilising a K-Nearest Neighbours (KNN) algorithm (KNNImputer, scikit-learn, Python v3.12) with inverse Euclidean distance weighting, using five neighbours for the whole cohort and three for the lymphomyeloid and PsA subsets. The complete feature matrix was then standardised utilising Z-score normalisation (StandardScaler) to scale all variables to a mean of 0 and a standard deviation of 1, so that variables with inherently large numerical ranges (e.g., digital cellularity) did not disproportionately dominate distance calculations during clustering.

*Unsupervised Hierarchical Clustering*

To define novel, data-driven disease pathotypes within the PsA cohort, unsupervised agglomerative hierarchical clustering was applied to the standardised feature matrix utilising the SciPy library (scipy.cluster.hierarchy). Linkage was computed via Ward's minimum variance method utilising a Euclidean distance metric. The whole cohort and the lymphomyeloid subset were clustered on the same features using k-means clustering (scikit-learn; 20 initialisations). Candidate numbers of clusters were evaluated using the Elbow method (within-cluster sum of squares) and average Silhouette scoring, calculated with k-means clustering. For the PsA cohort, three clusters were selected because three pathotypes were more informative than two or five and are analogous to the established RA pathotypes described by Humby et al.[8] The resulting phenotypic clusters were visualised alongside clinical treatment regimens and traditional pathotype classifications utilising annotated heatmaps, with features ordered by Ward hierarchical clustering.

*Visualisation of Longitudinal Treatment Trajectories*

To evaluate pathotype-specific therapeutic responses over time, longitudinal clinical data were visualised using swimmer plots. These plots mapped individual patient trajectories from the baseline index date (day of synovial biopsy) through sequential lines of therapy. Each discrete lane represented a single patient, with temporal annotations detailing the initiation, duration, and cessation of specific conventional synthetic or biologic DMARDs. Clinical response endpoints, defined by EULAR or PsARC criteria, were graphically overlaid onto these timelines. This visual framework facilitated the precise temporal analysis of treatment persistence, primary non-response, and secondary loss of efficacy, allowing for the direct correlation of longitudinal clinical outcomes with baseline synovial architecture.

*Development of the Clinical Diagnostic Algorithm*

To translate the high-dimensional, computationally derived clusters into a pragmatic tool for routine clinical histopathology, a stepwise diagnostic decision tree was formulated. Marker selection for this algorithm was driven by a combination of biological relevance and the statistical capacity to discriminate between the three novel PsA pathotypes (Dense Myeloid, Fibro-Stromal, and Angio-Lymphoid). This approach resulted in the selection of a focused four-marker panel (CD31, CD68, CD138, and Picrosirius Red) to be utilised alongside standard H&E.

The algorithm's structural architecture was developed iteratively through empirical testing of various semi-quantitative grading cut-offs (0–3) to balance classification accuracy with real-world clinical applicability. To ensure the workflow remained highly feasible and resource-efficient for standard pathology laboratories, the decision tree was strictly constrained to a maximum of three sequential evaluative steps per patient: commencing inherently with baseline H&E assessment, followed by the requisite evaluation of a maximum of two additional specialised stains to reach a final pathotype designation.

We acknowledge a methodological limitation inherent to this developmental phase: the optimisation of diagnostic grading thresholds utilising the identical dataset from which the underlying biological clusters were discovered introduces an inherent degree of circularity. Consequently, while the proposed algorithm accurately stratifies this specific discovery cohort and was explicitly designed around practical clinical constraints, its true diagnostic validity, predictive power, and generalisability must be robustly confirmed in an independent cohort.

**Supplementary Figure 1** Semi-quantitative scoring criteria. Criteria for scoring (a) CD3, CD20, CD68 and CD138, (b) CD31 and (c) picrosirius red with fast green.

**a**

Scoring criteria: CD3, CD20, CD68 and CD138

| Score | Description |
|---|---|
| 0 | Rare, scattered positive cells or complete absence of specific staining within the synovial sublining and lining layers. Background staining is clean, with no identifiable inflammatory aggregates or clusters. |
| 1 | Scattered positive cells distributed sparsely throughout the sublining tissue, or occasional very small, loose clusters comprising fewer than 10-20 positive cells. Staining is minimal and does not form cohesive inflammatory infiltrates. |
| 2 | Frequent positive cells forming distinct, multifocal aggregates or moderate-sized perivascular cuffs within the sublining. Infiltrates are prominent across multiple high-power fields but do not form dense, confluent sheets. |
| 3 | Dense, confluent sheets of positive cells, or large, well-organized ectopic lymphoid-like aggregates spanning extensive areas of the synovial sublining. Infiltration is heavy and widespread throughout the tissue section. |

**b**

Scoring criteria: CD31 vascularity

| Score | Description |
|---|---|
| 0 | Minimal, baseline vascular staining with rare or absent distinct endothelial profiles identifiable within the synovial tissue. |
| 1 | Faint or background staining with a few scattered, well-defined CD31-positive vessels sparsely distributed within the sublining layer. |
| 2 | Clear, obvious endothelial staining with several distinct blood vessels visible across the sublining layer, reflecting an intermediate level of vascularity. |
| 3 | Prominent, heavy staining with numerous CD31-positive vessels densely populating throughout the sublining layer, characteristic of marked synovial angiogenesis. |

**c**

Scoring criteria: Picrosirius red with fast green fibrosis

| Score | Description |
|---|---|
| 0 | Complete absence of red collagen staining, with the tissue displaying only background counterstain (fast green) without detectable fibrosis. |
| 1 | Limited, focal deposition of collagen, with red staining observed in less than 25% of the synovial tissue area. |
| 2 | Clear, prominent fibrosis with red collagen deposition distributed across at least half of the synovial tissue section. |
| 3 | Extensive, heavy fibrosis with more than half, and up to the majority, of the synovial tissue occupied by dense red, dark red, or intensely birefringent collagen networks. |

**Supplementary Figure 2** Representative images of immunohistochemistry and histochemistry scores. Examples of consensus semi-quantitative scores from 0 to 3 for each stain. Images are shown at different magnifications.

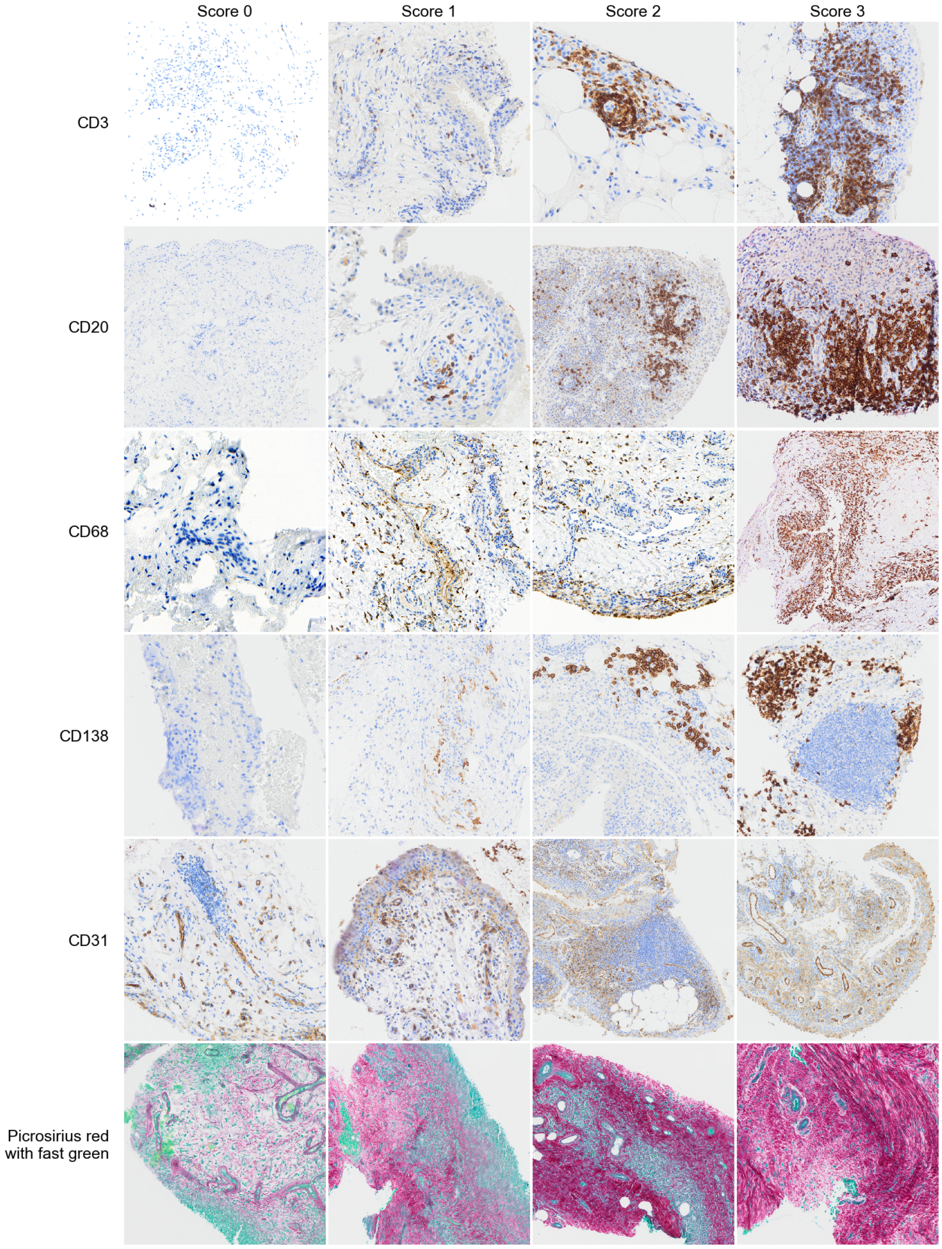

**Supplementary Figure 3** Representative images of Krenn scores from 0–9. Examples of total Krenn scores from 0 to 9, with the corresponding grade. H&E. Some sections are also shown in Figure 1e, because one section can illustrate more than one score. No section in this cohort scored 9, so the example shown for a score of 9 is illustrative. Images are shown at different magnifications.

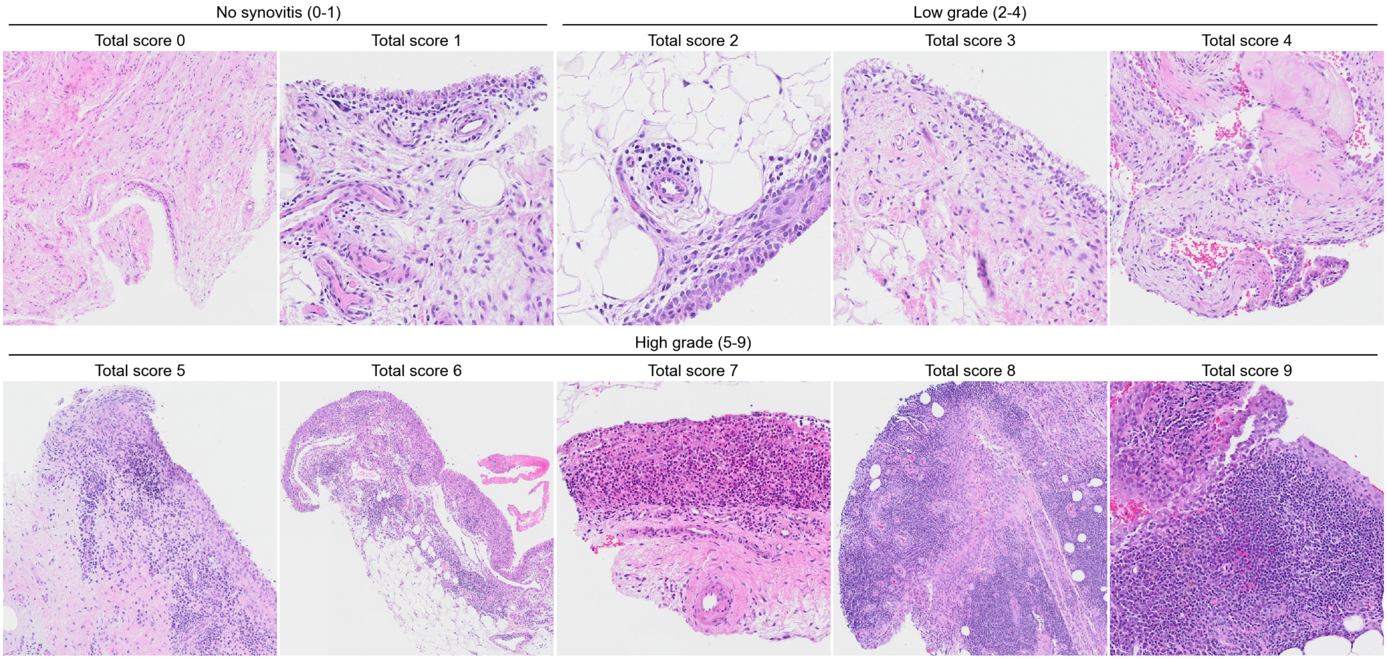

**Supplementary Figure 4** Representative images of pathotypes in RA. Representative sections for each pathotype in (a) seropositive RA and (b) seronegative RA. One patient per pathotype. Images are shown at different magnifications.

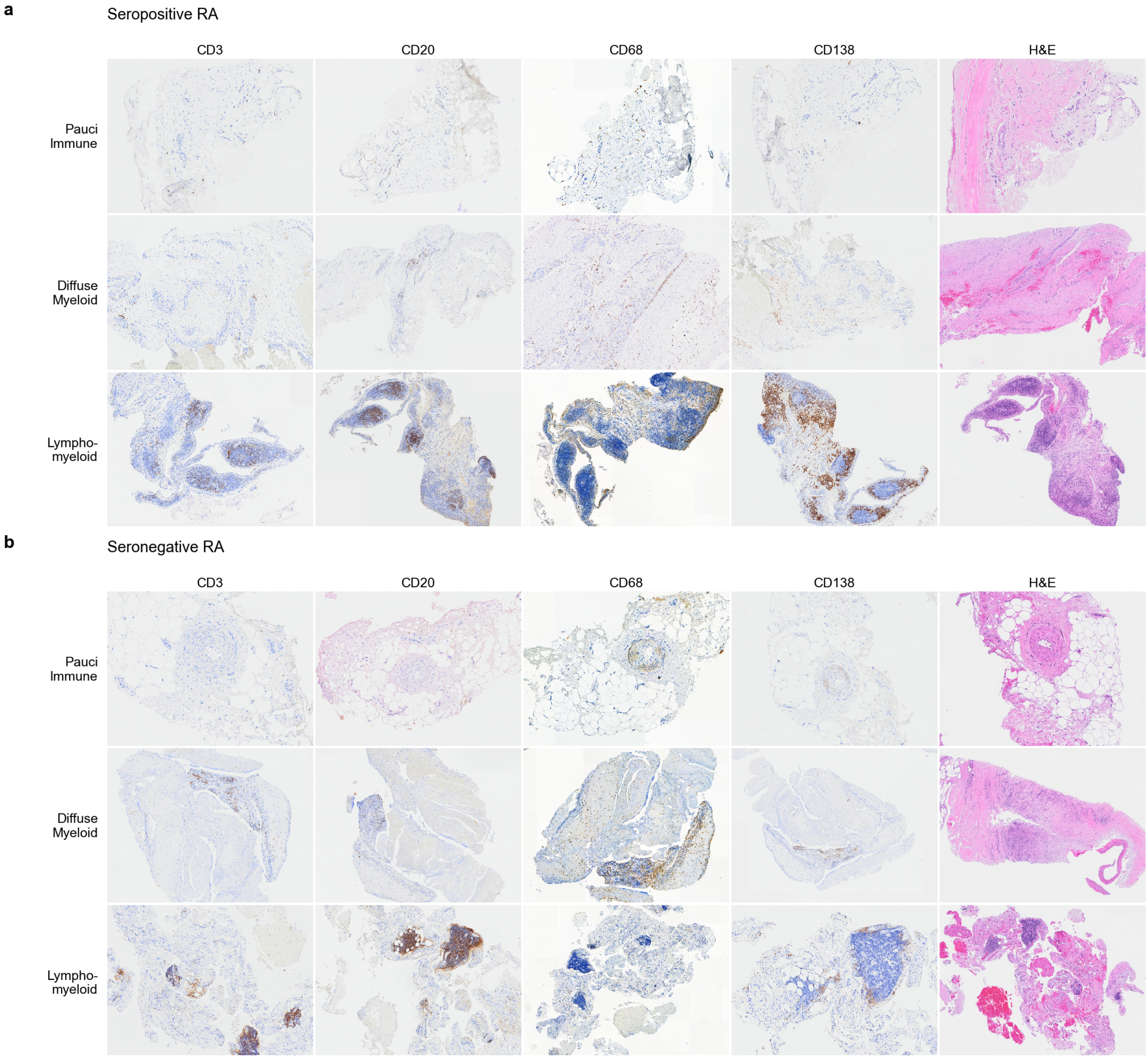

**Supplementary Figure 5** Krenn synovitis scores and components show significant correlation with several other histological markers. (a, b) Synovial pathotype by diagnosis in patients with (a) low-grade and (b) high-grade synovitis. Numbers within bars are patients. (c) Spearman correlations between Krenn scores and histological markers. *P<0.05, **P<0.01, ***P<0.001, unadjusted.

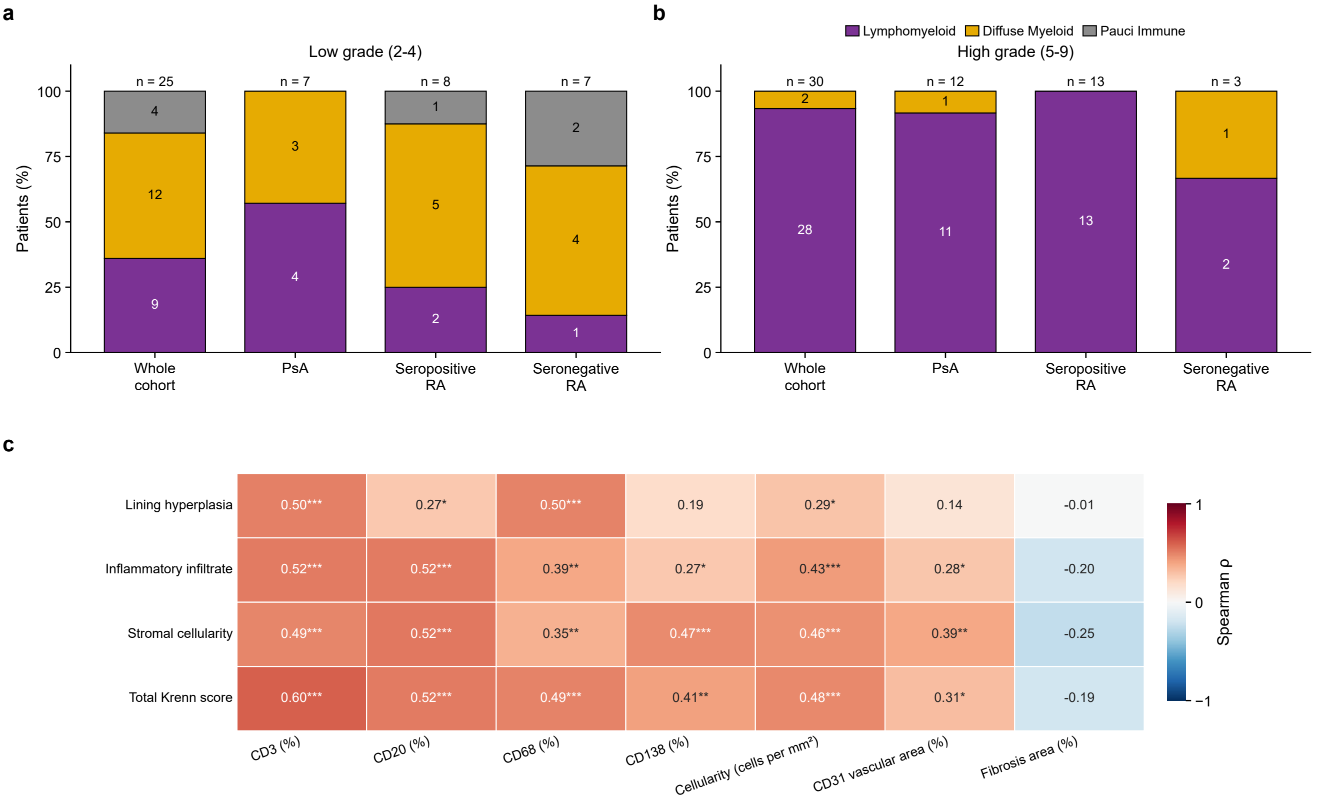

**Supplementary Figure 6** Digital pathology scores show concordance with consensus manual semi-quantitative scores. (a–d) Digital percentage of positive cells by consensus manual score for CD3, CD20, CD68 and CD138, with Spearman's $\rho$. Boxes show the median and IQR, whiskers extend to 1.5 × IQR, and points are individual patients. (e–h) ROC curves showing cut-offs for the digital pathology scores that align with a consensus semi-quantitative score of $\geq 2$. Points mark the cut-off with the highest Youden's J, and the dashed line shows chance. Cut-offs were chosen and evaluated in the same sections.

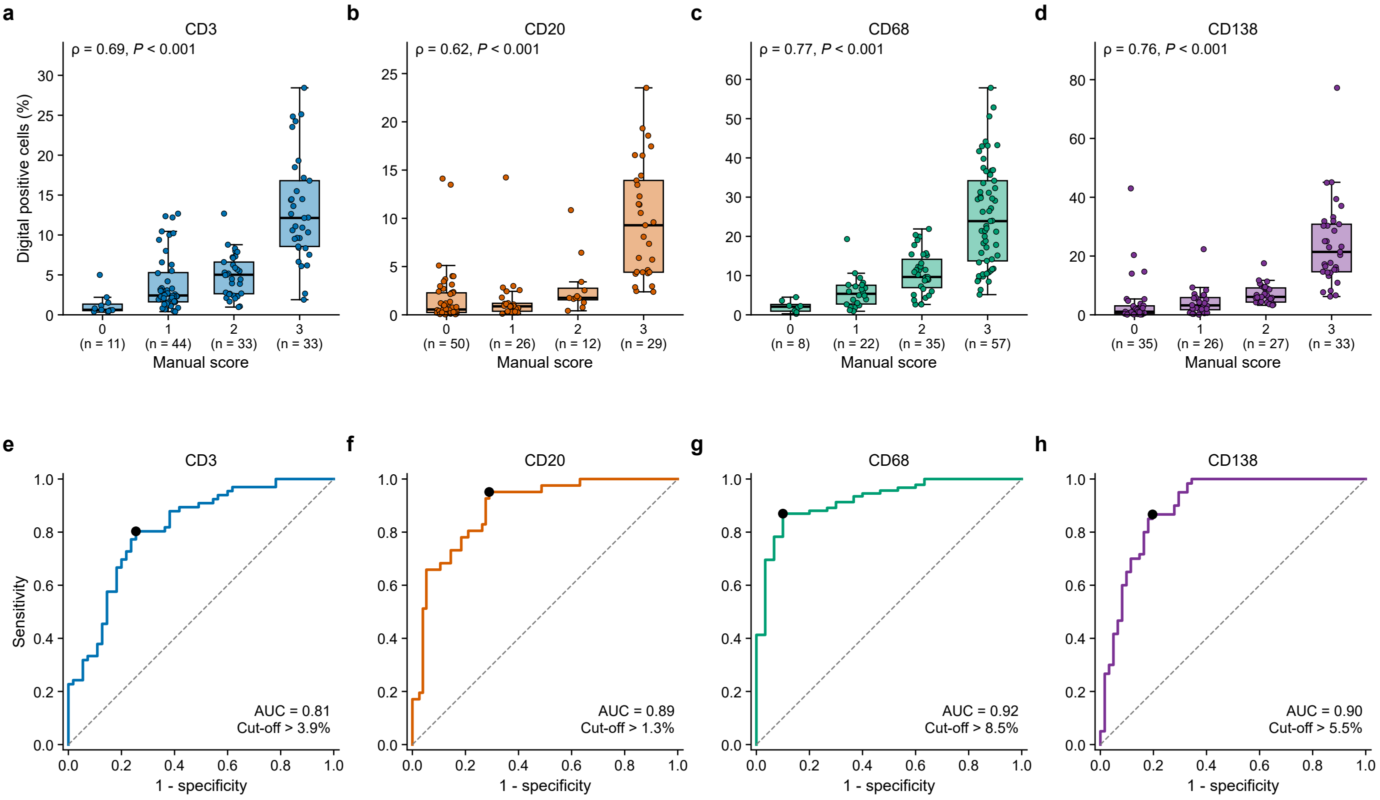

**Digital thresholds for a consensus semi-quantitative score of $\geq 2$**

| Stain | N Blocks | Spearman Rho | AUC | Digital Threshold | Sensitivity | Specificity |
|---|---|---|---|---|---|---|
| CD3 | 121 | 0.69 | 0.813 | > 3.88% | 80.3% | 74.5% |
| CD20 | 117 | 0.618 | 0.888 | > 1.28% | 95.1% | 71.1% |
| CD68 | 122 | 0.766 | 0.921 | > 8.47% | 87.0% | 90.0% |
| CD138 | 121 | 0.756 | 0.896 | > 5.46% | 86.7% | 80.3% |

**Supplementary Figure 7** Histological markers by diagnosis. (a–d) Maximum consensus semi-quantitative score per patient for CD3, CD20, CD68 and CD138 by diagnosis. Boxes show the median and IQR, whiskers extend to 1.5 × IQR, and points are individual patients. (e) Heatmap of median digital positivity by diagnosis, with post-hoc comparison of CD68 digital positivity. Brackets mark false discovery rate-adjusted pairwise comparisons among PsA, seropositive RA and seronegative RA only (**q<0.01). Other IA, other inflammatory arthritis.

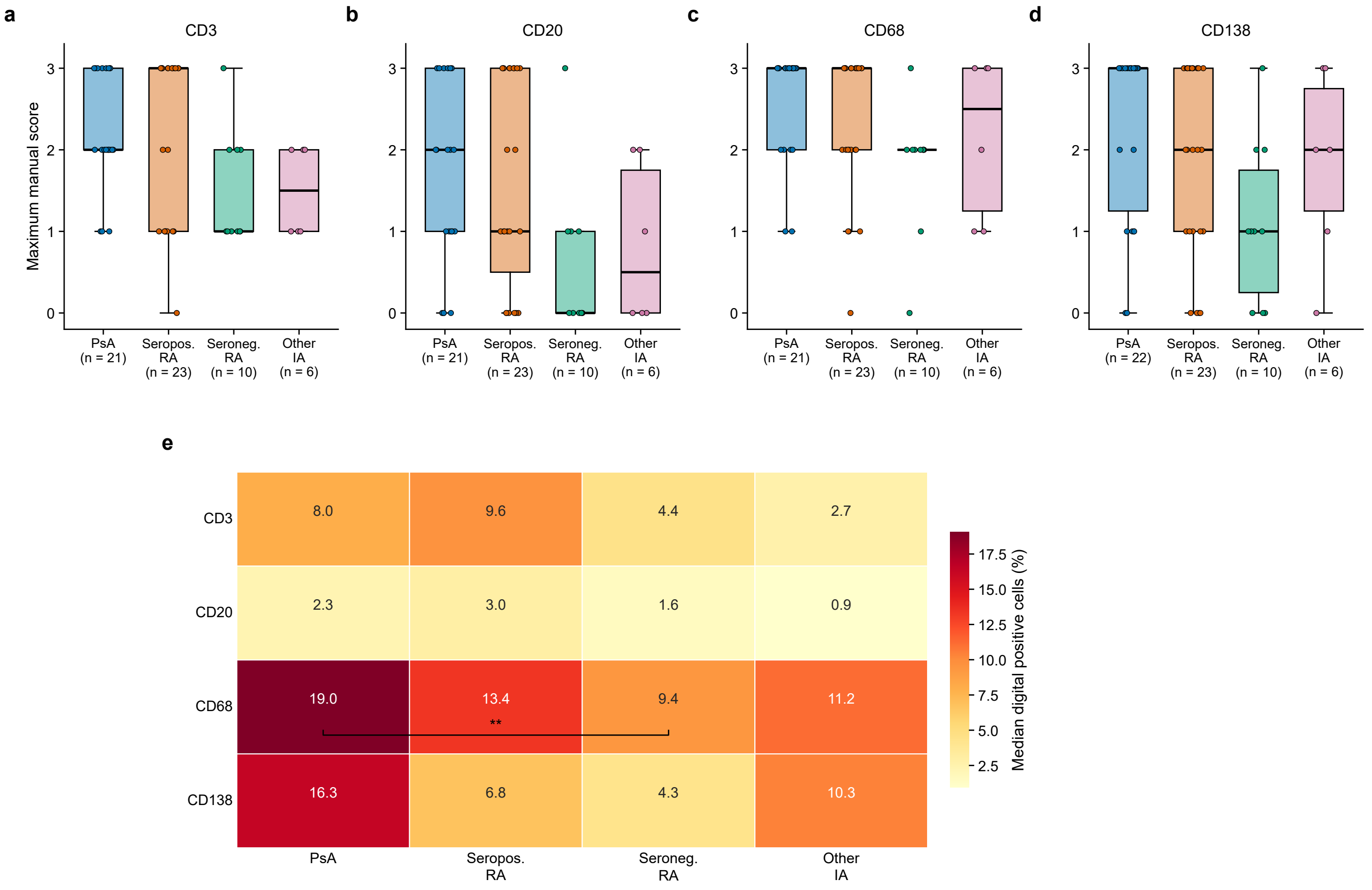

**Supplementary Figure 8** Histological markers by treatment at biopsy. (a) Heatmap of median histological markers by treatment at biopsy. Colour shows the median scaled within each row, and numbers are the medians. (b) Maximum cellularity by treatment at biopsy, with Kruskal-Wallis P value. Boxes show the median and IQR, whiskers extend to 1.5 × IQR, and points are individual patients. csDMARD, conventional synthetic disease-modifying antirheumatic drug; b/tsDMARD, biologic or targeted synthetic DMARD.

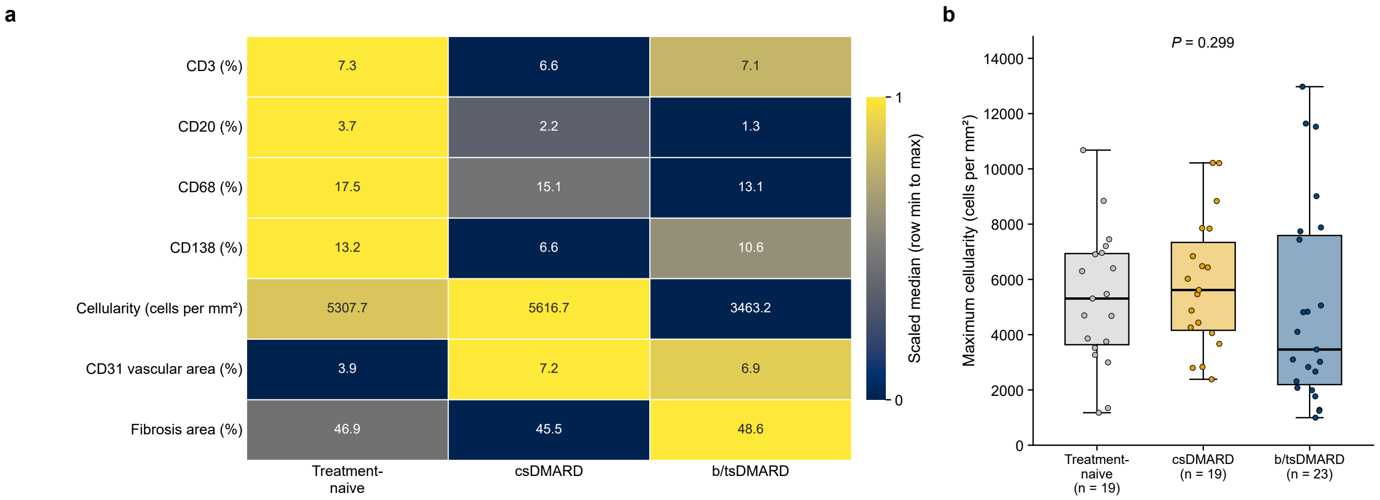

**Supplementary Figure 9** Disease duration shows no correlation with histological markers or pathotypes. (a–g) Scatterplots of disease duration and histological markers with Spearman rank coefficients. Lines show least-squares fits with 95% confidence bands. (h) Pathotype distribution by disease duration. Numbers within bars are patients. The table shows multivariable regression modelling the independent effect of disease duration on histological markers, adjusted for age, sex, BMI, treatment status and biologic status.

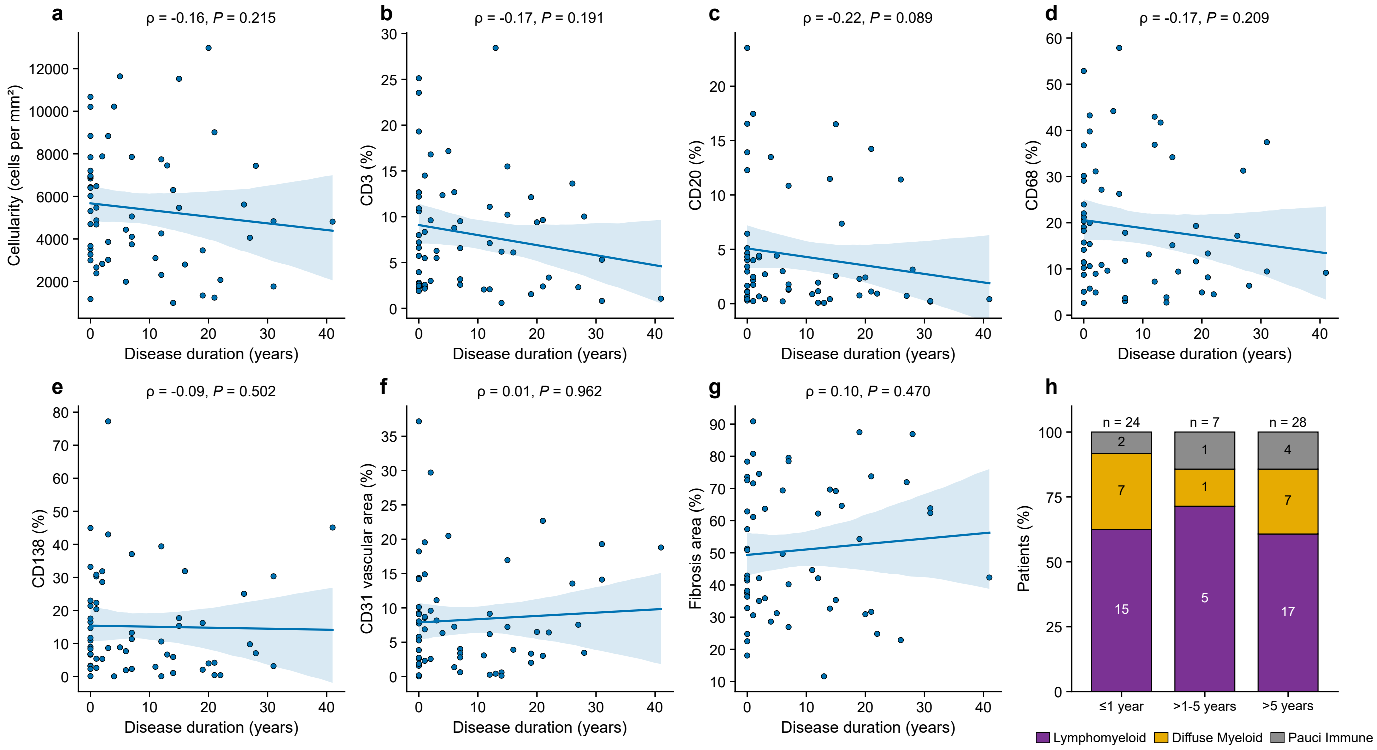

##### Multivariable regression of histological markers on disease duration

| Histological marker | N | Coefficient (per year) | 95% CI | p-value | q-value (BH) |
|---|---|---|---|---|---|
| Cellularity | 60 | −33.309 | −122.181 to 55.562 | 0.456 | 0.797 |
| CD3 | 59 | −0.104 | −0.294 to 0.086 | 0.278 | 0.797 |
| CD20 | 59 | 0.002 | −0.162 to 0.166 | 0.981 | 0.981 |
| CD68 | 59 | −0.195 | −0.634 to 0.244 | 0.377 | 0.797 |
| CD138 | 60 | 0.031 | −0.420 to 0.481 | 0.892 | 0.981 |
| VAF | 59 | 0.017 | −0.224 to 0.258 | 0.887 | 0.981 |
| Fibrosis | 57 | 0.349 | −0.318 to 1.016 | 0.298 | 0.797 |

**Supplementary Figure 10** Unsupervised clustering of the entire cohort does not show clear clustering by diagnosis or RA pathotype, suggesting similarities between PsA and RA synovial inflammation. (a) Elbow method and (b) silhouette analysis for candidate numbers of clusters; three clusters were used (dashed line). (c) Heatmap of synovial histological markers across the cohort, with patients grouped by k-means cluster (k=3). The tables show the composition and median histological markers of each cluster.

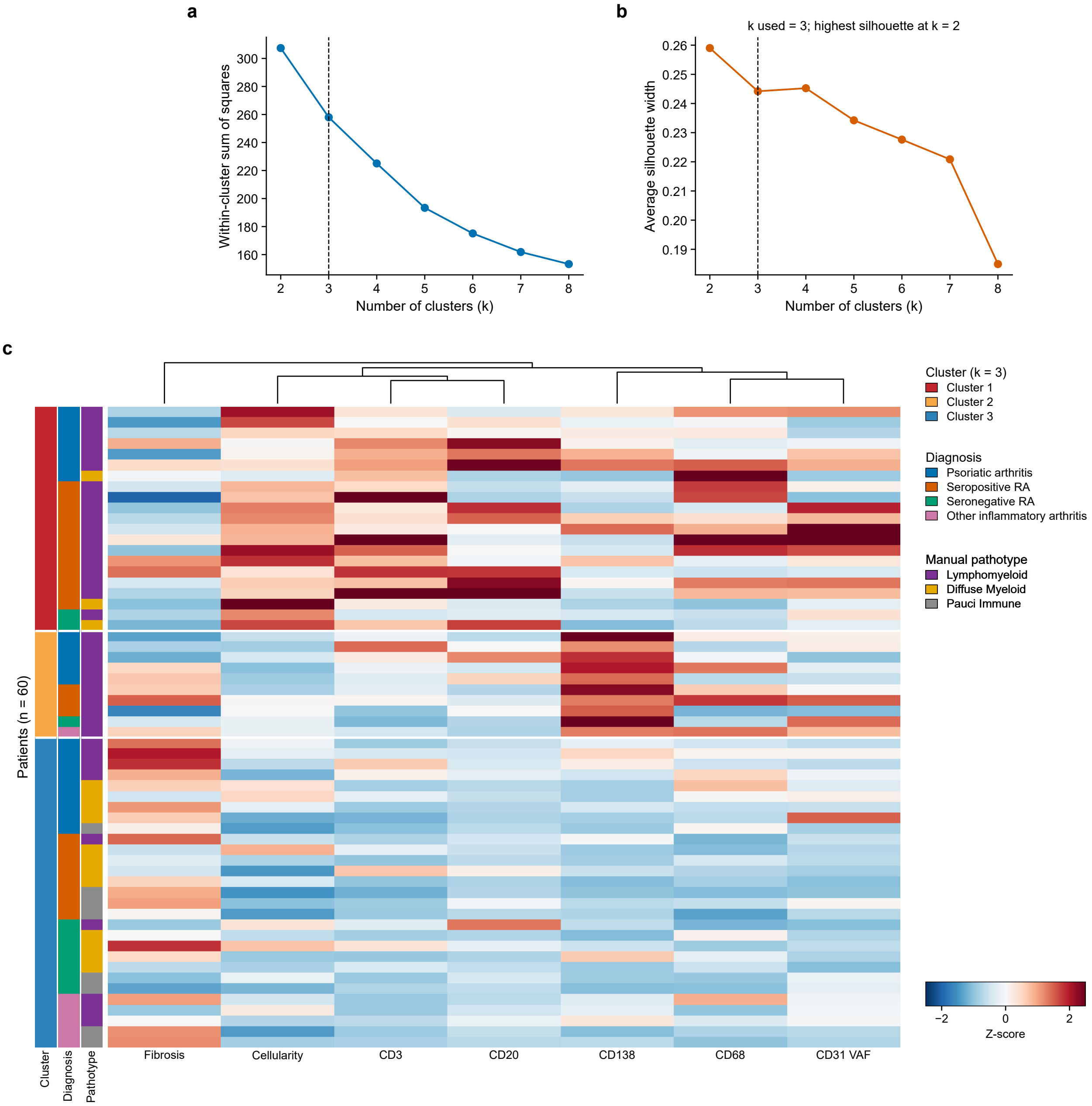

#### Cluster composition

| Cluster | N | Cohort % | PsA (n) | Sero-positive RA (n) | Sero-negative RA (n) | Other IA (n) | Lympho-myeloid (n) | Diffuse Myeloid (n) | Pauci Immune (n) |
|---|---|---|---|---|---|---|---|---|---|
| Cluster 1 | 21 | 35.0% | 7 | 12 | 2 | 0 | 18 | 3 | 0 |
| Cluster 2 | 10 | 16.7% | 5 | 3 | 1 | 1 | 10 | 0 | 0 |
| Cluster 3 | 29 | 48.3% | 9 | 8 | 7 | 5 | 9 | 12 | 8 |

#### Median histological markers (maximum per patient)

| Cluster | N | Cohort % | CD3 (%) | CD20 (%) | CD68 (%) | CD138 (%) | Cellular-ity (/mm$^2$) | VAF (%) | Fibrosis (%) |
|---|---|---|---|---|---|---|---|---|---|
| Cluster 1 | 21 | 35.0% | 12.64 | 4.64 | 23.97 | 10.60 | 7838.2 | 11.12 | 36.91 |
| Cluster 2 | 10 | 16.7% | 6.70 | 2.20 | 19.51 | 35.13 | 3886.4 | 8.70 | 42.32 |
| Cluster 3 | 29 | 48.3% | 2.68 | 1.12 | 10.24 | 5.35 | 3463.2 | 3.42 | 62.19 |

**Supplementary Figure 11** Unsupervised clustering of the lymphomyeloid pathotype cohort shows a distinct cluster of PsA patients separate from seropositive RA, suggesting a disease-specific subpathotype. (a) Elbow method and (b) silhouette analysis for candidate numbers of clusters; two clusters were used (dashed line). (c) Heatmap of synovial histological markers across the lymphomyeloid pathotype cohort, with patients grouped by k-means cluster (k=2). The tables show the composition and median histological markers of each cluster.

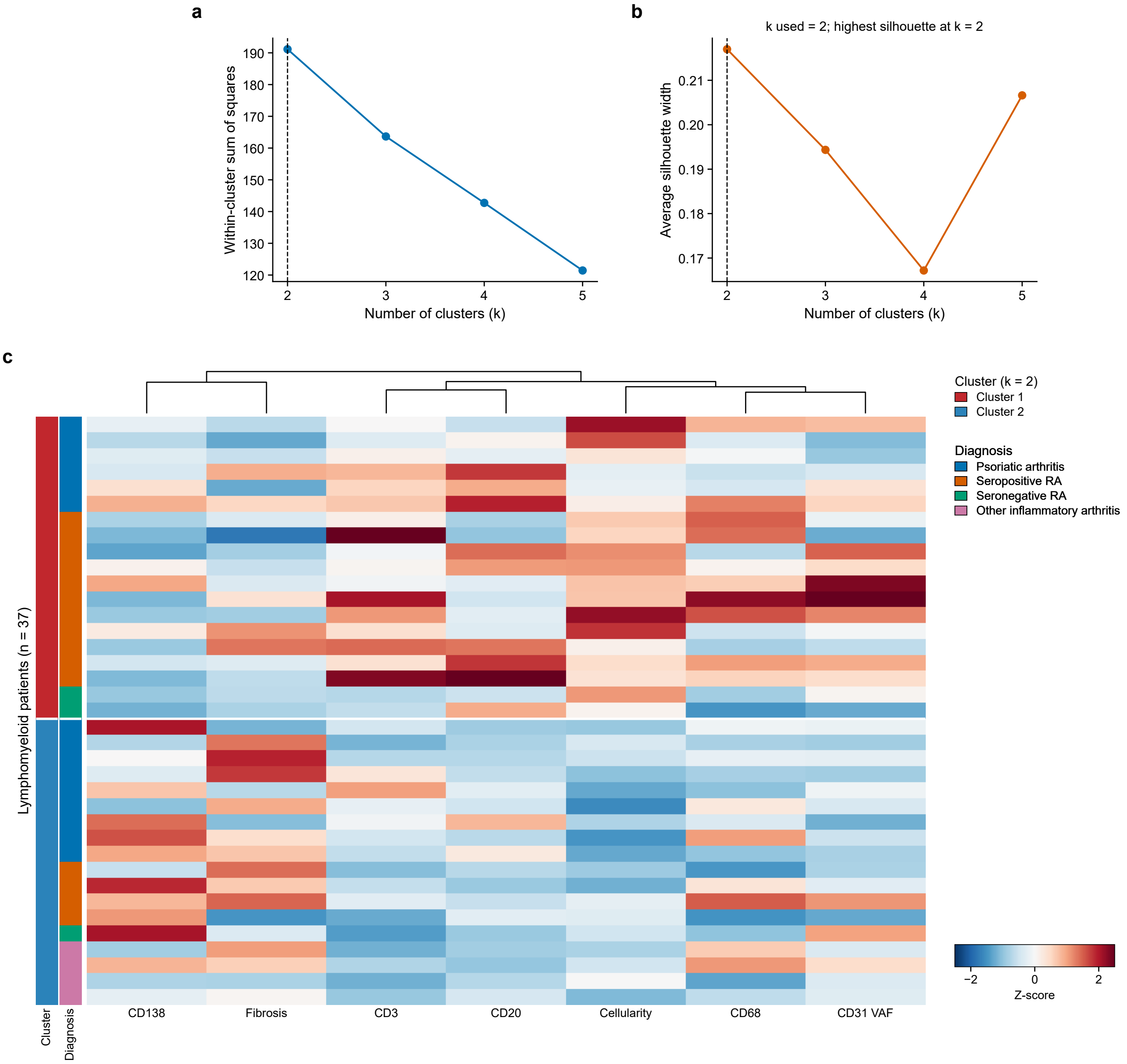

#### Cluster composition

| Cluster | N | Subset % | PsA (n) | Seropositive RA (n) | Seronegative RA (n) | Other IA (n) |
|---|---|---|---|---|---|---|
| Cluster 1 | 19 | 51.4% | 6 | 11 | 2 | 0 |
| Cluster 2 | 18 | 48.6% | 9 | 4 | 1 | 4 |

#### Median histological markers (maximum per patient)

| Cluster | N | Subset % | CD3 (%) | CD20 (%) | CD68 (%) | CD138 (%) | Cellularity (/mm$^2$) | VAF (%) | Fibrosis (%) |
|---|---|---|---|---|---|---|---|---|---|
| Cluster 1 | 19 | 51.4% | 12.64 | 6.44 | 23.97 | 11.73 | 7739.7 | 13.55 | 36.91 |
| Cluster 2 | 18 | 48.6% | 5.79 | 1.85 | 15.99 | 29.45 | 3902.0 | 7.64 | 63.68 |

**Supplementary Figure 12** Unsupervised hierarchical clustering of the PsA cohort shows three clusters, suggesting novel PsA pathotypes. (a) Elbow method and (b) silhouette analysis for candidate numbers of clusters; three clusters were used (dashed line). (c) Heatmap of synovial histological markers across the PsA cohort, with patients grouped by Ward hierarchical cluster (k=3). The tables show the treatment, manual pathotype and median histological markers of each novel pathotype.

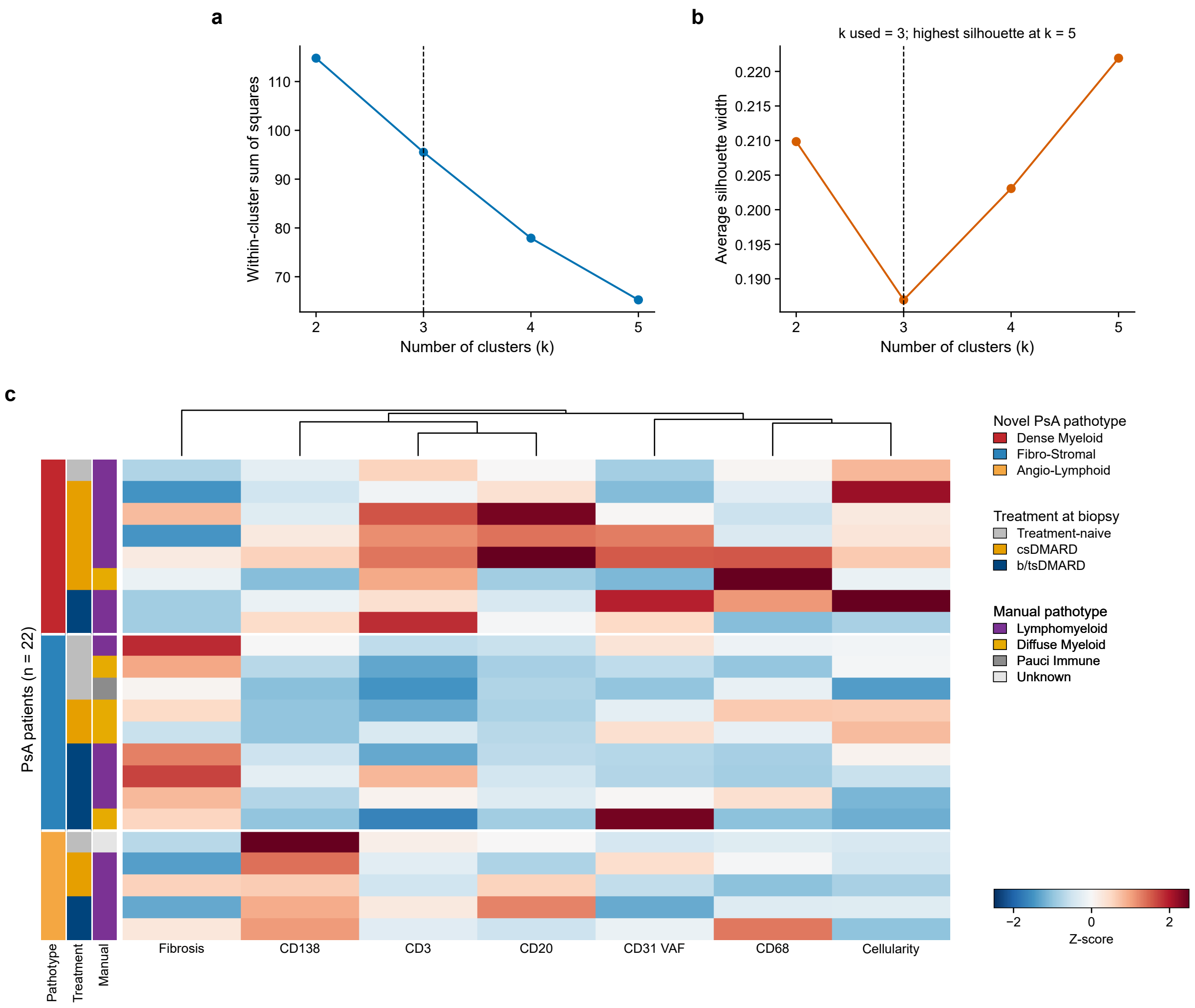

#### Treatment at biopsy and manual pathotype by novel pathotype

| Cluster | N | Cohort % | Treatment naive (n) | csDMARD (n) | bD-MARD/ tsDMARD (n) | Lympho-myeloid (n) | Diffuse Myeloid (n) | Pauci Immune (n) |
|---|---|---|---|---|---|---|---|---|
| Dense Myeloid | 8 | 36.4% | 1 | 5 | 2 | 7 | 1 | 0 |
| Fibro-Stromal | 9 | 40.9% | 3 | 2 | 4 | 4 | 4 | 1 |
| Angio-Lymphoid | 5 | 22.7% | 1 | 2 | 2 | 4 | 0 | 0 |

#### Median histological markers (maximum per patient)

| Cluster | N | Cohort % | CD3 (%) | CD20 (%) | CD68 (%) | CD138 (%) | Cellular-ity ($/mm^2$) | VAF (%) | Fibrosis (%) |
|---|---|---|---|---|---|---|---|---|---|
| Dense Myeloid | 8 | 36.4% | 13.15 | 5.54 | 20.14 | 17.08 | 6048.1 | 8.41 | 35.32 |
| Fibro-Stromal | 9 | 40.9% | 2.79 | 1.13 | 19.05 | 7.68 | 4680.0 | 5.79 | 69.38 |
| Angio-Lymphoid | 5 | 22.7% | 7.16 | 4.65 | 19.51 | 39.39 | 3669.2 | 5.04 | 26.92 |

**Supplementary Figure 13** Boxplots of clinical disease activity measures by novel PsA pathotype. (a) Tender joint count, (b) swollen joint count, (c) patient global score, (d) physician global score, (e) psoriasis body surface area, (f) ESR and (g) CRP. Boxes show the median and IQR, whiskers extend to 1.5 × IQR, and points are individual patients. Kruskal-Wallis P values are unadjusted and exploratory.

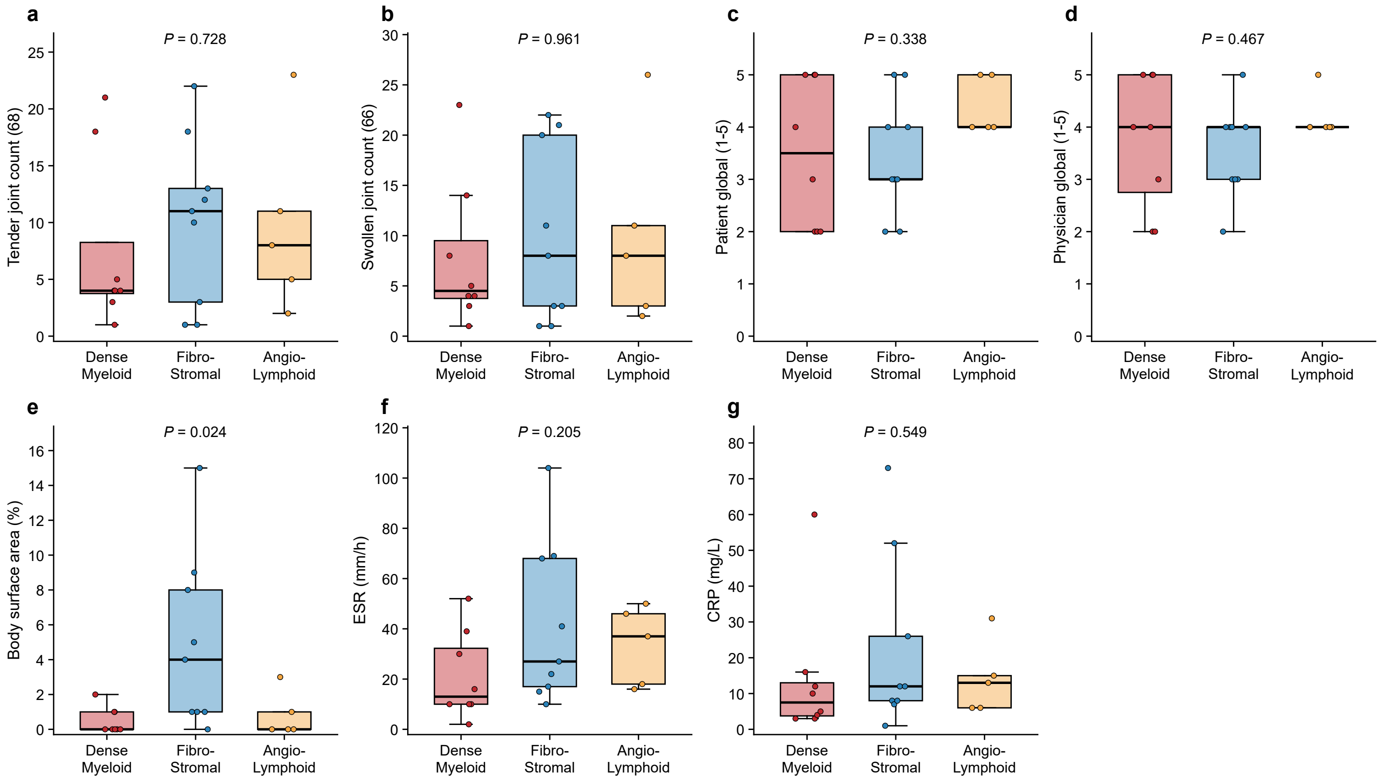

**Supplementary Figure 14** Scatterplots of histological markers vs clinical disease activity measures. Correlations between digital histology and clinical measures in PsA with unadjusted P<0.05. $\rho$, Spearman correlation. Lines show least-squares fits with 95% confidence bands, clipped to the observed range.

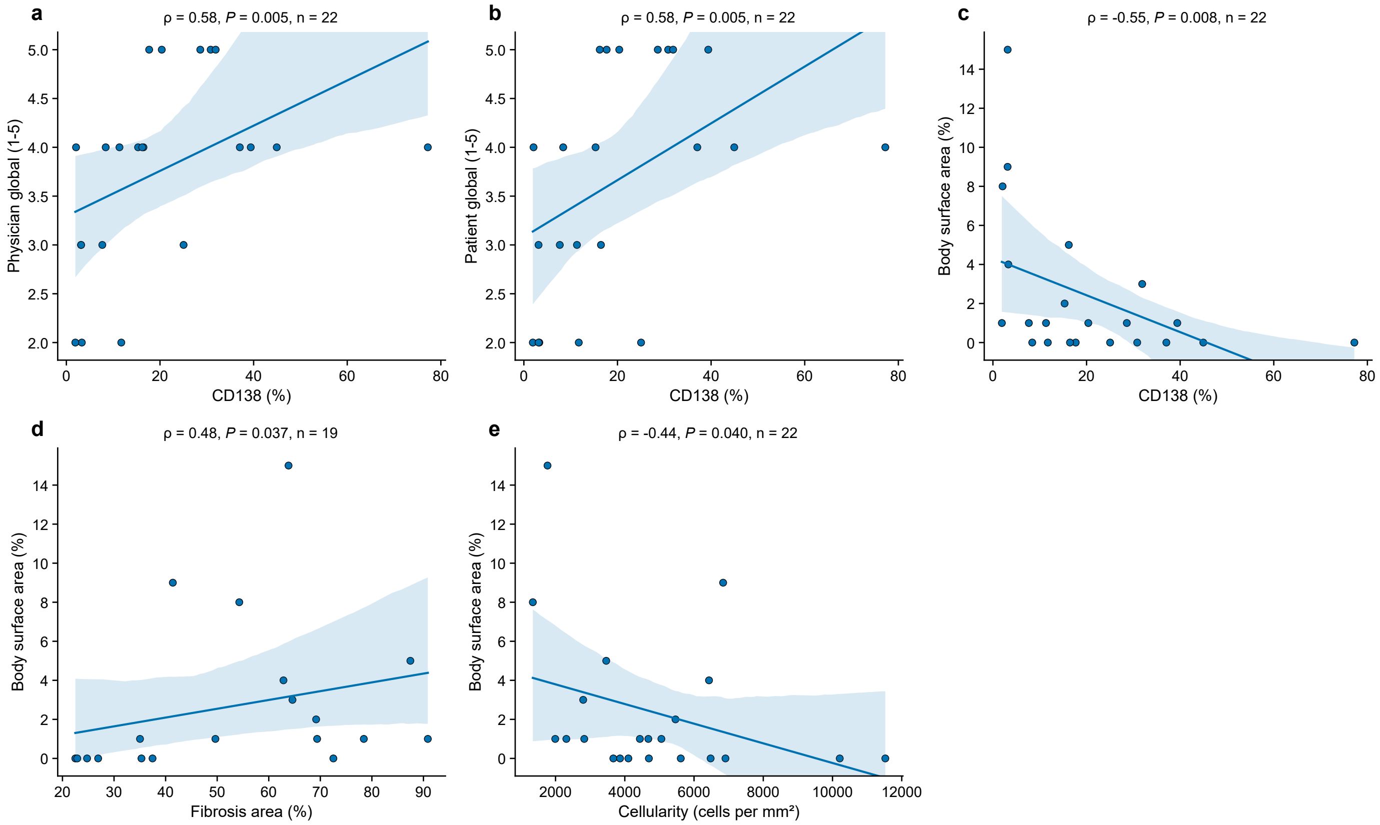

**Supplementary Figure 15** Treatment response outcomes and medication changes by treatment at biopsy for the entire cohort. (a) Swimmer plot of longitudinal treatment response outcomes and (b) swimmer plot of longitudinal medication changes, stratified by treatment at biopsy. Each row is one patient, and rows are in the same order in both panels.

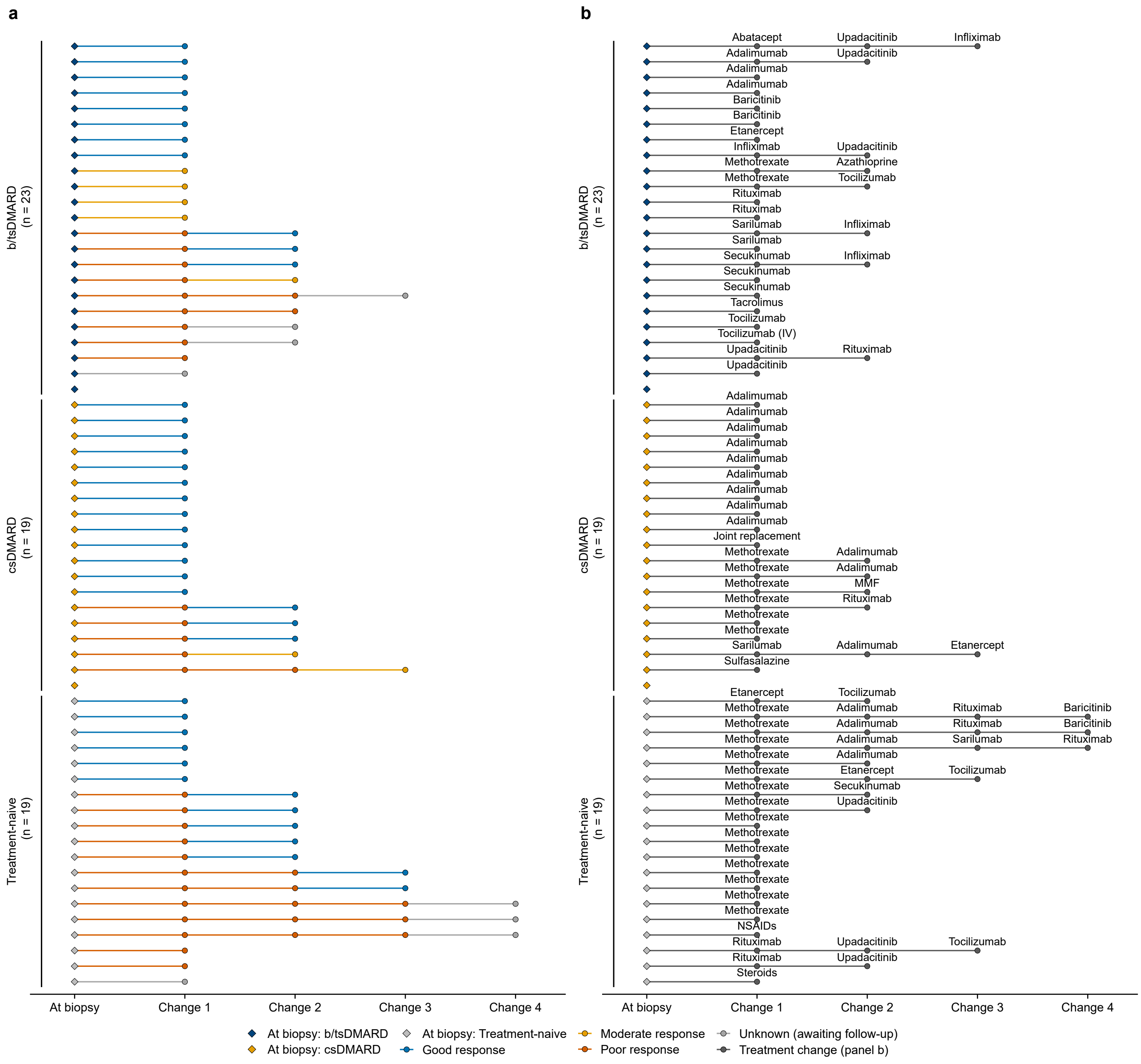

**Supplementary Figure 16** Treatment response outcomes and medication changes by treatment at biopsy for the PsA cohort, and follow-up duration. (a) Swimmer plot of longitudinal treatment response outcomes and (b) swimmer plot of longitudinal medication changes for the PsA cohort, stratified by treatment at biopsy. Each row is one patient, and rows are in the same order in both panels. (c) Follow-up duration in years by diagnosis. Boxes show the median and IQR, whiskers extend to 1.5 × IQR, and points are individual patients. Groups with fewer than three patients are shown as points only.

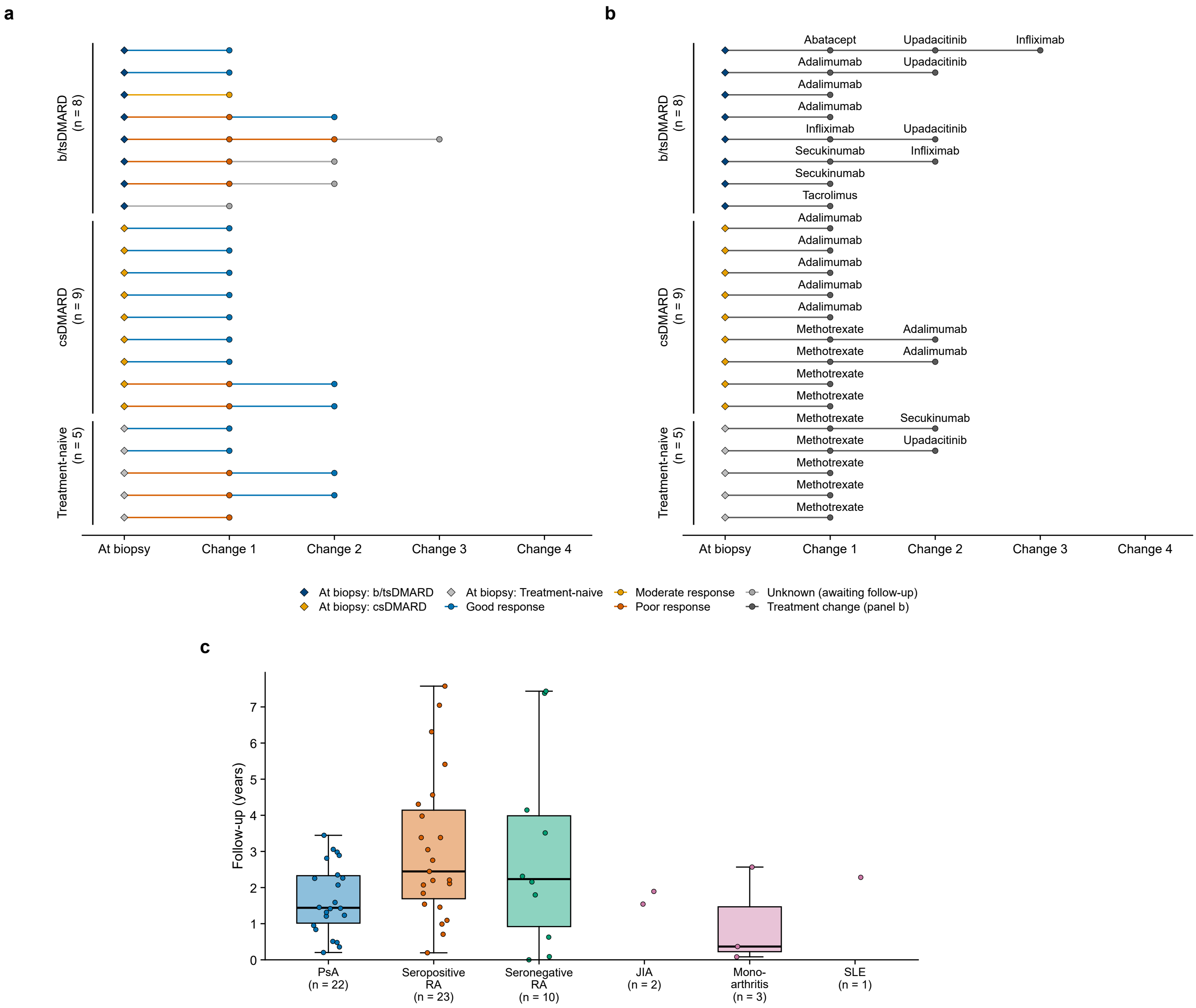

**Supplementary Figure 17** Concurrent OA in the PsA cohort, and joint biopsied. (a–d) OA severity as determined by knee radiographs for the PsA cohort (n=22): (a) overall, and by (b) novel PsA synovial pathotype, (c) patient age and (d) disease duration. Patients without a radiograph are shown as not available. (e) Joint biopsied in the entire cohort (n=61) and (f) by diagnosis. Numbers are patients.

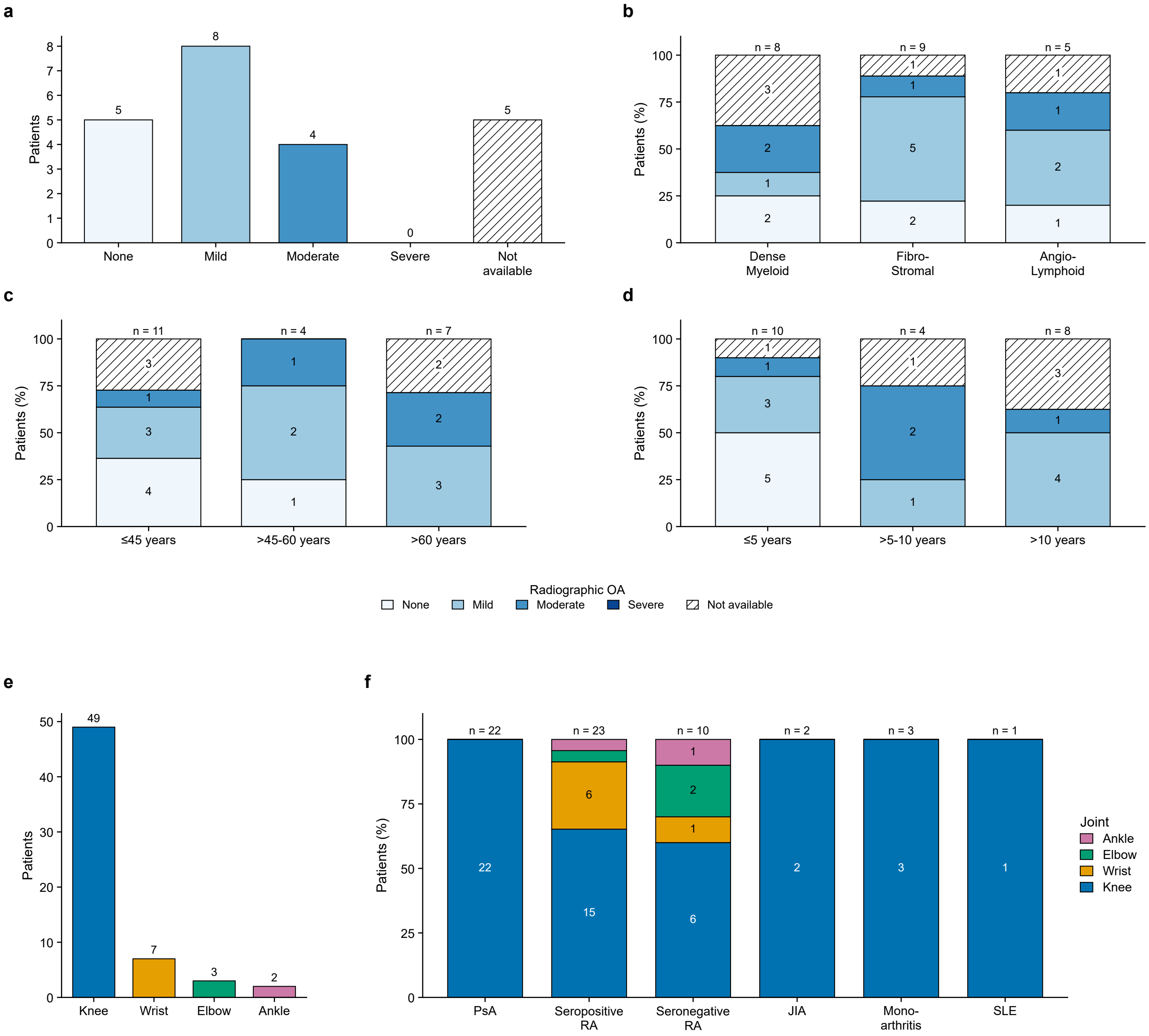

**Supplementary Table 1** Krenn synovitis scores by domain and overall grading by diagnosis

| Domain | Entire Cohort (n=61) | PsA (n=22) | Seropositive RA (n=23) | Sero-negative RA (n=10) | p-value (PsA vs Seropositive RA) | p-value (PsA vs Sero-negative RA) | p-value (Sero-positive RA vs Sero-negative RA) |
|---|---|---|---|---|---|---|---|
| Lining Hyperplasia (Median [IQR]) | 1.0 (1.0–2.0) | 1.5 (1.0–2.0) | 1.0 (1.0–2.0) | 1.0 (1.0–1.0) | 0.904 | 0.157 | 0.107 |
| Inflammatory Infiltrate (Median [IQR]) | 1.0 (1.0–2.0) | 1.0 (1.0–2.0) | 2.0 (1.0–2.0) | 1.0 (1.0–2.0) | 0.763 | 0.695 | 0.561 |
| Stromal Cellularity (Median [IQR]) | 1.0 (1.0–3.0) | 2.0 (1.0–2.0) | 2.0 (1.0–3.0) | 1.0 (1.0–1.0) | 0.611 | 0.113 | 0.101 |
| Total Krenn Score (Median [IQR]) | 4.0 (3.0–6.0) | 5.0 (3.0–6.0) | 5.0 (3.0–6.0) | 2.5 (2.0–4.8) | 0.863 | 0.144 | 0.132 |
| **Total Krenn Grade (n [%])** | | | | | | | |
| No synovitis (0–1) | 5 (8.2%) | 2 (9.1%) | 2 (8.7%) | 0 (0.0%) | 0.991 | 0.178 | 0.151 |
| Low grade (2–4) | 26 (42.6%) | 8 (36.4%) | 8 (34.8%) | 7 (70.0%) | | | |
| High grade (5–9) | 30 (49.2%) | 12 (54.5%) | 13 (56.5%) | 3 (30.0%) | | | |

**Supplementary Table 2** Immunohistochemistry marker digital positivity by diagnosis

| Stain | N (Patients) | PsA Median (IQR) | Seropositive RA Median (IQR) | Seronegative RA Median (IQR) | Kruskal-Wallis Statistic | p-value |
|---|---|---|---|---|---|---|
| CD3 | 60 | 8.0 (5.5–12.1) | 9.6 (2.9–12.7) | 4.4 (2.4–6.1) | 3.781 | 0.151 |
| CD20 | 60 | 2.3 (0.8–6.4) | 3.0 (1.0–4.8) | 1.6 (0.5–3.0) | 0.929 | 0.629 |
| CD68 | 60 | 19.0 (11.8–26.3) | 13.4 (6.1–33.9) | 9.4 (5.9–11.3) | 6.995 | 0.030 |
| CD138 | 61 | 16.3 (7.9–30.3) | 6.8 (4.2–18.0) | 4.3 (0.9–8.2) | 6.016 | 0.049 |

**Supplementary Table 3** Vascularity and fibrosis scores by diagnosis

| Metric | Entire Cohort (n=60) | PsA (n=21) | Seropositive RA (n=23) | Sero-negative RA (n=10) | p-value (PsA vs Seropositive RA) | p-value (PsA vs Sero-negative RA) | p-value (Sero-positive RA vs Sero-negative RA) |
|---|---|---|---|---|---|---|---|
| CD31 Vascular Area Fraction (%) (Median [IQR]) | 6.50 (2.76–10.62) | 6.18 (3.34–9.24) | 8.36 (2.86–17.26) | 4.90 (2.37–6.76) | 0.474 | 0.410 | 0.200 |
| Picrosirius Red Fibrosis (%) (Median [IQR]) | 47.15 (35.11–69.33) | 54.29 (35.18–69.28) | 42.91 (36.92–66.66) | 37.04 (31.10–49.04) | 0.800 | 0.302 | 0.248 |

**Supplementary Table 4** Correlation between clinical measures of disease activity in PsA by novel pathotype

| Clinical Variable | N (Patients) | Dense Myeloid Median (IQR) | Fibro-Stromal Median (IQR) | Angio-Lymphoid Median (IQR) | Kruskal-Wallis Statistic | p-value |
|---|---|---|---|---|---|---|
| Tender Joint Count (68) | 22 | 4.0 (3.8–8.2) | 11.0 (3.0–13.0) | 8.0 (5.0–11.0) | 0.635 | 0.728 |
| Swollen Joint Count (66) | 22 | 4.5 (3.8–9.5) | 8.0 (3.0–20.0) | 8.0 (3.0–11.0) | 0.080 | 0.961 |
| Patient Global (1–5) | 22 | 3.5 (2.0–5.0) | 3.0 (3.0–4.0) | 4.0 (4.0–5.0) | 2.171 | 0.338 |
| Physician Global (1–5) | 22 | 4.0 (2.8–5.0) | 4.0 (3.0–4.0) | 4.0 (4.0–4.0) | 1.524 | 0.467 |
| Body Surface Area (BSA) | 22 | 0.0 (0.0–1.0) | 4.0 (1.0–8.0) | 0.0 (0.0–1.0) | 7.467 | 0.024 |
| ESR | 22 | 13.0 (10.0–32.2) | 27.0 (17.0–68.0) | 37.0 (18.0–46.0) | 3.169 | 0.205 |
| CRP | 22 | 7.5 (3.8–13.0) | 12.0 (8.0–26.0) | 13.0 (6.0–15.0) | 1.198 | 0.549 |

**Supplementary Table 5** Correlation between histological markers and clinical measures of disease activity in PsA

| **Histological Marker** | **Clinical Measure** | **N** | **Spearman r** | **p-value** |
|---|---|---|---|---|
| CD138 (%) | Physician Global | 22 | 0.581 | 0.0046 |
| CD138 (%) | Patient Global | 22 | 0.577 | 0.0049 |
| CD138 (%) | Body Surface Area (BSA) | 22 | −0.551 | 0.0079 |
| Fibrosis (%) | Body Surface Area (BSA) | 19 | 0.481 | 0.0370 |
| Cellularity (cells/mm$^2$) | Body Surface Area (BSA) | 22 | −0.440 | 0.0403 |
| Cellularity (cells/mm$^2$) | ESR | 22 | −0.379 | 0.0815 |
| CD20 (%) | Body Surface Area (BSA) | 21 | −0.384 | 0.0857 |
| CD3 (%) | Body Surface Area (BSA) | 21 | −0.379 | 0.0906 |
| CD20 (%) | Physician Global | 21 | 0.363 | 0.1053 |
| CD3 (%) | ESR | 21 | −0.363 | 0.1058 |
| CD20 (%) | ESR | 21 | −0.332 | 0.1418 |
| CD20 (%) | Patient Global | 21 | 0.301 | 0.1852 |
| CD68 (%) | Tender Joint Count (68) | 21 | −0.272 | 0.2321 |
| Vascular Area Fraction (%) | Physician Global | 21 | 0.272 | 0.2334 |
| Fibrosis (%) | Patient Global | 19 | 0.287 | 0.2335 |
| Vascular Area Fraction (%) | ESR | 21 | 0.271 | 0.2346 |
| CD68 (%) | Body Surface Area (BSA) | 21 | −0.269 | 0.2383 |
| Cellularity (cells/mm$^2$) | CRP | 22 | −0.247 | 0.2673 |
| CD20 (%) | Swollen Joint Count (66) | 21 | 0.239 | 0.2963 |
| Cellularity (cells/mm$^2$) | Swollen Joint Count (66) | 22 | 0.229 | 0.3056 |
| CD3 (%) | CRP | 21 | −0.221 | 0.3351 |
| Cellularity (cells/mm$^2$) | Patient Global | 22 | −0.214 | 0.3400 |
| Fibrosis (%) | Physician Global | 19 | 0.224 | 0.3565 |
| CD68 (%) | ESR | 21 | −0.205 | 0.3720 |
| Vascular Area Fraction (%) | Patient Global | 21 | 0.190 | 0.4100 |
| CD3 (%) | Patient Global | 21 | 0.188 | 0.4151 |
| Vascular Area Fraction (%) | CRP | 21 | 0.184 | 0.4242 |
| CD68 (%) | Swollen Joint Count (66) | 21 | −0.182 | 0.4288 |
| CD3 (%) | Tender Joint Count (68) | 21 | −0.173 | 0.4540 |
| CD68 (%) | Physician Global | 21 | −0.162 | 0.4842 |
| CD3 (%) | Physician Global | 21 | 0.159 | 0.4899 |
| Cellularity (cells/mm$^2$) | Tender Joint Count (68) | 22 | 0.139 | 0.5378 |
| CD20 (%) | Tender Joint Count (68) | 21 | 0.137 | 0.5540 |
| Fibrosis (%) | ESR | 19 | −0.143 | 0.5584 |
| CD138 (%) | Swollen Joint Count (66) | 22 | 0.129 | 0.5658 |
| Vascular Area Fraction (%) | Swollen Joint Count (66) | 21 | −0.122 | 0.5996 |
| CD138 (%) | CRP | 22 | 0.109 | 0.6286 |
| CD20 (%) | CRP | 21 | −0.107 | 0.6452 |
| Vascular Area Fraction (%) | Tender Joint Count (68) | 21 | −0.104 | 0.6528 |
| CD68 (%) | CRP | 21 | −0.096 | 0.6779 |
| Fibrosis (%) | Swollen Joint Count (66) | 19 | −0.094 | 0.7009 |
| Cellularity (cells/mm$^2$) | Physician Global | 22 | −0.083 | 0.7144 |
| CD3 (%) | Swollen Joint Count (66) | 21 | −0.075 | 0.7482 |
| CD138 (%) | Tender Joint Count (68) | 22 | 0.065 | 0.7752 |
| CD68 (%) | Patient Global | 21 | −0.065 | 0.7809 |
| Fibrosis (%) | Tender Joint Count (68) | 19 | 0.046 | 0.8524 |
| Fibrosis (%) | CRP | 19 | 0.030 | 0.9034 |
| Vascular Area Fraction (%) | Body Surface Area (BSA) | 21 | 0.026 | 0.9120 |
| CD138 (%) | ESR | 22 | 0.016 | 0.9442 |

**Supplementary Table 6** Follow-up duration by diagnosis

| Group | N (Patients) | Median Follow-up [IQR] (Years) | Range (Years) |
|---|---|---|---|
| Whole Cohort (All Diagnoses) | 61 | 2.11 [1.21–3.05] | 0.00–7.58 |
| Diagnosis: Juvenile Idiopathic Arthritis | 2 | 1.72 [1.63–1.81] | 1.54–1.89 |
| Diagnosis: Monoarthritis | 3 | 0.37 [0.22–1.47] | 0.08–2.57 |
| Diagnosis: PsA | 22 | 1.44 [1.02–2.33] | 0.20–3.45 |
| Diagnosis: Seronegative RA | 10 | 2.23 [0.92–3.99] | 0.00–7.44 |
| Diagnosis: Seropositive RA | 23 | 2.45 [1.69–4.14] | 0.19–7.58 |
| Diagnosis: Systemic Lupus Erythematosus | 1 | 2.28 [2.28–2.28] | 2.28–2.28 |

**Supplementary Table 7** Treatment changes post biopsy by diagnosis

| Treatment Change | Entire Cohort (n=59) | PsA (n=22) | Seropositive RA (n=23) | Seronegative RA (n=9) | JIA (n=2) | Mono-arthritis (n=2) | SLE (n=1) |
|---|---|---|---|---|---|---|---|
| Methotrexate | 22 (37.3%) | 9 (40.9%) | 9 (39.1%) | 2 (22.2%) | 1 (50.0%) | 0 (0.0%) | 1 (100.0%) |
| Adalimumab | 12 (20.3%) | 8 (36.4%) | 3 (13.0%) | 1 (11.1%) | 0 (0.0%) | 0 (0.0%) | 0 (0.0%) |
| Rituximab | 4 (6.8%) | 0 (0.0%) | 4 (17.4%) | 0 (0.0%) | 0 (0.0%) | 0 (0.0%) | 0 (0.0%) |
| Secukinumab | 3 (5.1%) | 2 (9.1%) | 0 (0.0%) | 0 (0.0%) | 1 (50.0%) | 0 (0.0%) | 0 (0.0%) |
| Sarilumab | 3 (5.1%) | 0 (0.0%) | 1 (4.3%) | 2 (22.2%) | 0 (0.0%) | 0 (0.0%) | 0 (0.0%) |
| Etanercept | 2 (3.4%) | 0 (0.0%) | 0 (0.0%) | 2 (22.2%) | 0 (0.0%) | 0 (0.0%) | 0 (0.0%) |
| Upadacitinib | 2 (3.4%) | 0 (0.0%) | 2 (8.7%) | 0 (0.0%) | 0 (0.0%) | 0 (0.0%) | 0 (0.0%) |
| Baricitinib | 2 (3.4%) | 0 (0.0%) | 2 (8.7%) | 0 (0.0%) | 0 (0.0%) | 0 (0.0%) | 0 (0.0%) |
| NSAIDs | 1 (1.7%) | 0 (0.0%) | 0 (0.0%) | 0 (0.0%) | 0 (0.0%) | 1 (50.0%) | 0 (0.0%) |
| Tacrolimus | 1 (1.7%) | 1 (4.5%) | 0 (0.0%) | 0 (0.0%) | 0 (0.0%) | 0 (0.0%) | 0 (0.0%) |
| Tocilizumab | 1 (1.7%) | 0 (0.0%) | 1 (4.3%) | 0 (0.0%) | 0 (0.0%) | 0 (0.0%) | 0 (0.0%) |
| Abatacept | 1 (1.7%) | 1 (4.5%) | 0 (0.0%) | 0 (0.0%) | 0 (0.0%) | 0 (0.0%) | 0 (0.0%) |
| Tocilizumab (SC to IV) | 1 (1.7%) | 0 (0.0%) | 1 (4.3%) | 0 (0.0%) | 0 (0.0%) | 0 (0.0%) | 0 (0.0%) |
| Sulfasalazine | 1 (1.7%) | 0 (0.0%) | 0 (0.0%) | 1 (11.1%) | 0 (0.0%) | 0 (0.0%) | 0 (0.0%) |
| Joint Replacement | 1 (1.7%) | 0 (0.0%) | 0 (0.0%) | 0 (0.0%) | 0 (0.0%) | 1 (50.0%) | 0 (0.0%) |
| Steroids | 1 (1.7%) | 0 (0.0%) | 0 (0.0%) | 1 (11.1%) | 0 (0.0%) | 0 (0.0%) | 0 (0.0%) |
| Infliximab | 1 (1.7%) | 1 (4.5%) | 0 (0.0%) | 0 (0.0%) | 0 (0.0%) | 0 (0.0%) | 0 (0.0%) |

**Supplementary Table 8** Longitudinal treatment response outcomes for the PsA cohort by novel pathotype

| Treatment Stage | Total PsA | p-value (Efficacy) | Dense Myeloid (n=8) | Fibro-Stromal (n=9) | Angio-Lymphoid (n=5) |
|---|---|---|---|---|---|
| **Treatment Change 1 (Total n=22)** | | | [Stage n=8] | [Stage n=9] | [Stage n=5] |
| Good Response | 11 (50.0%) | 0.396 | 4 (50.0%) | 4 (44.4%) | 3 (60.0%) |
| Moderate Response | 1 (4.5%) | | 0 (0.0%) | 0 (0.0%) | 1 (20.0%) |
| Poor Response | 9 (40.9%) | | 4 (50.0%) | 4 (44.4%) | 1 (20.0%) |
| Unknown (Awaiting FU) | 1 (4.5%) | | 0 (0.0%) | 1 (11.1%) | 0 (0.0%) |
| **Treatment Change 2 (Total n=8)** | | | [Stage n=4] | [Stage n=3] | [Stage n=1] |
| Good Response | 5 (62.5%) | 1.000 | 2 (50.0%) | 3 (100.0%) | 0 (0.0%) |
| Poor Response | 1 (12.5%) | | 1 (25.0%) | 0 (0.0%) | 0 (0.0%) |
| Unknown (Awaiting FU) | 2 (25.0%) | | 1 (25.0%) | 0 (0.0%) | 1 (100.0%) |
| **Treatment Change 3 (Total n=1)** | | | [Stage n=1] | [Stage n=0] | [Stage n=0] |
| Unknown (Awaiting FU) | 1 (100.0%) | | 1 (100.0%) | 0 (0.0%) | 0 (0.0%) |
